# Pre-existing polymerase-specific T cells expand in abortive seronegative SARS-CoV-2 infection

**DOI:** 10.1101/2021.06.26.21259239

**Authors:** Leo Swadling, Mariana O. Diniz, Nathalie M. Schmidt, Oliver E. Amin, Aneesh Chandran, Emily Shaw, Corinna Pade, Joseph M. Gibbons, Nina Le Bert, Anthony T. Tan, Anna Jeffery-Smith, Cedric Tan, Christine Y. L. Tham, Stephanie Kucyowicz, Gloryanne Aidoo-Micah, Joshua Rosenheim, Jessica Davies, Melanie P. Jensen, George Joy, Laura E McCoy, Ana M Valdes, Lucy van Dorp, Daniel M. Altmann, Rosemary J. Boyton, Charlotte Manisty, Thomas A. Treibel, James C. Moon, COVIDsortium investigators, Francois Balloux, Áine McKnight, Mahdad Noursadeghi, Antonio Bertoletti, Mala K. Maini

## Abstract

Individuals with likely exposure to the highly infectious SARS-CoV-2 do not necessarily develop PCR or antibody positivity, suggesting some may clear sub-clinical infection before seroconversion. T cells can contribute to the rapid clearance of SARS-CoV-2 and other coronavirus infections^1–5^. We hypothesised that pre-existing memory T cell responses, with cross-protective potential against SARS-CoV-2^6–12^, would expand *in vivo* to mediate rapid viral control, potentially aborting infection. We studied T cells against the replication transcription complex (RTC) of SARS-CoV-2 since this is transcribed first in the viral life cycle^13–15^and should be highly conserved. We measured SARS-CoV-2-reactive T cells in a cohort of intensively monitored healthcare workers (HCW) who remained repeatedly negative by PCR, antibody binding, and neutralisation for SARS-CoV-2 (exposed seronegative, ES). 16-weeks post-recruitment, ES had memory T cells that were stronger and more multispecific than an unexposed pre-pandemic cohort, and more frequently directed against the RTC than the structural protein-dominated responses seen post-detectable infection (matched concurrent cohort). The postulate that HCW with the strongest RTC-specific T cells had an abortive infection was supported by a low-level increase in IFI27 transcript, a robust early innate signature of SARS-CoV-2 infection^16^. We showed that the RNA-polymerase within RTC was the largest region of high sequence conservation across human seasonal coronaviruses (HCoV) and was preferentially targeted by T cells from UK and Singapore pre-pandemic cohorts and from ES. RTC epitope-specific T cells capable of cross-recognising HCoV variants were identified in ES. Longitudinal samples from ES and an additional validation cohort, showed pre-existing RNA-polymerase-specific T cells expanded *in vivo* following SARS-CoV-2 exposure, becoming enriched in the memory response of those with abortive compared to overt infection. In summary, we provide evidence of abortive seronegative SARS-CoV-2 infection with expansion of cross-reactive RTC-specific T cells, highlighting these highly conserved proteins as targets for future vaccines against endemic and emerging *Coronaviridae*.

## Main

There is wide variability in the outcome of exposure to highly infectious SARS-CoV-2, ranging from severe illness to asymptomatic infection, to those remaining negative with standard diagnostic tests. Identification of exposure without infection has largely been based on isolated cases with single time-point screening^6,17–20^. We undertook a systematic study of an intensively monitored cohort of healthcare workers (HCW) exposed during the first pandemic wave, comparing those with or without PCR and/or antibody evidence of SARS-CoV-2 infection. We postulated that in HCW where PCR, and the most sensitive binding and neutralising antibody (nAb) tests, remained repeatedly negative (exposed seronegative, ES), T cell assays might distinguish a subset with a subclinical, rapidly terminated (abortive) infection. We hypothesised that these individuals would exhibit pre-existing memory T cells with cross-reactive potential, obviating the time required for *de novo* T cell priming and clonal expansion.

Recent studies have identified SARS-CoV-2 T cell reactivity in pre-pandemic samples^7,8,25,9–12,21–24^or exposed individuals who have not seroconverted^6,17–21^. However, the rapid global spread of SARS-CoV-2 has limited opportunities to follow individuals pre- and post-exposure to distinguish pre-existing T cells and their capacity to respond to SARS-CoV-2 infection *in vivo*. In ES HCW, and an additionally recruited cohort of medical students and laboratory staff with stored pre-pandemic samples that remained seronegative after close contact with cases, we had the unique opportunity to compare SARS-CoV-2-specific memory T cells with those already present in the same individual before, or at the time of, likely exposure.

Most studies have focused on T cells directed against SARS-CoV-2 structural proteins, with few analysing those against the large open reading frame (ORF)1^25^. We included analysis of T cells directed against the RTC within ORF1ab (RNA-polymerase co-factor NSP7, RNA-polymerase NSP12, and helicase NSP13) as putative targets for pre-existing responses with pan-*Coronaviridae* reactivity because they are likely to be highly conserved due to their key roles in the viral lifecycle. Consistent with this, where immunity against other viruses (including dengue, HBV, HCV and HIV) has been described in exposed seronegative individuals^26–29^, T cells were noted to be more likely to target non-structural viral proteins, such as polymerase, than in those with seropositive infection^30–35^. In the case of positive single-stranded RNA viruses like SARS-CoV-2, the 5’ end containing the RTC is immediately transcribed upon entering the cytoplasm, before the generation of subgenomic RNA templates for structural proteins^13,14^. Hence RTC antigens from SARS-CoV-2 should also be the first encountered by any pre-existing responses that could mediate early viral control.

### SARS-CoV-2 T cell responses in exposed seronegative HCW

We examined T cell reactivity in a subset of intensively monitored HCW from the COVIDsortium who did not develop documented SARS-CoV-2 infection despite likely exposure during the first UK pandemic wave. These were compared with HCW matched for exposure risk and demographic factors, who did develop evidence of laboratory-confirmed SARS-CoV-2 infection by PCR and/or seroconversion **(Fig. 1a;** Demographics **Extended Data Table 1)**. Additional control cohorts comprised healthy adults sampled in two geographical locations (London, UK; Singapore) prior to SARS-CoV-2 circulation in humans (pre-pandemic cohort; **Fig. 1a**). The exposed seronegative (ES) HCW group were defined by negativity on state-of-the-art diagnostic tests carried out weekly for 16weeks (SARS-CoV-2 PCR from nasopharyngeal swabs; anti-Spike-1 IgG and anti-nucleoprotein (NP) IgG/IgM seroassays^36^**(Fig. 1b-d)**. Having previously reported a range of nAb titres persisting at wk16 in the laboratory-confirmed infection group^21^, we examined nAb in the ES group. Two HCW with nAb titres just above the threshold were excluded from further analyses; the remaining ES were negative by pseudotype assay **(Fig. 1e)**, with a subset also confirmed negative at 3 time points by authentic virus neutralisation assay **(Extended Data Fig. 1a)**. Some may have been infected before recruitment but non-seroconverters after PCR positivity were rare (only 2.6% of PCR+ HCW were negative by all 3 serological tests^21^), and antibody responses were unlikely to have waned before study recruitment^36^. Furthermore staining ES with dual-colour tetramers showed they lacked detectable SARS-CoV-2 spike-specific memory B cells, which we have shown persist after waning of nAb^37^(example plots **Extended Data Fig. 1b**, comparable frequency to pre-pandemics [below the threshold of detection^37^], **Fig. 1f**), Thus, ES represented a cohort of intensely monitored HCW who resisted classical laboratory-confirmed infection during the first pandemic wave in the UK.

**Fig. 1.**
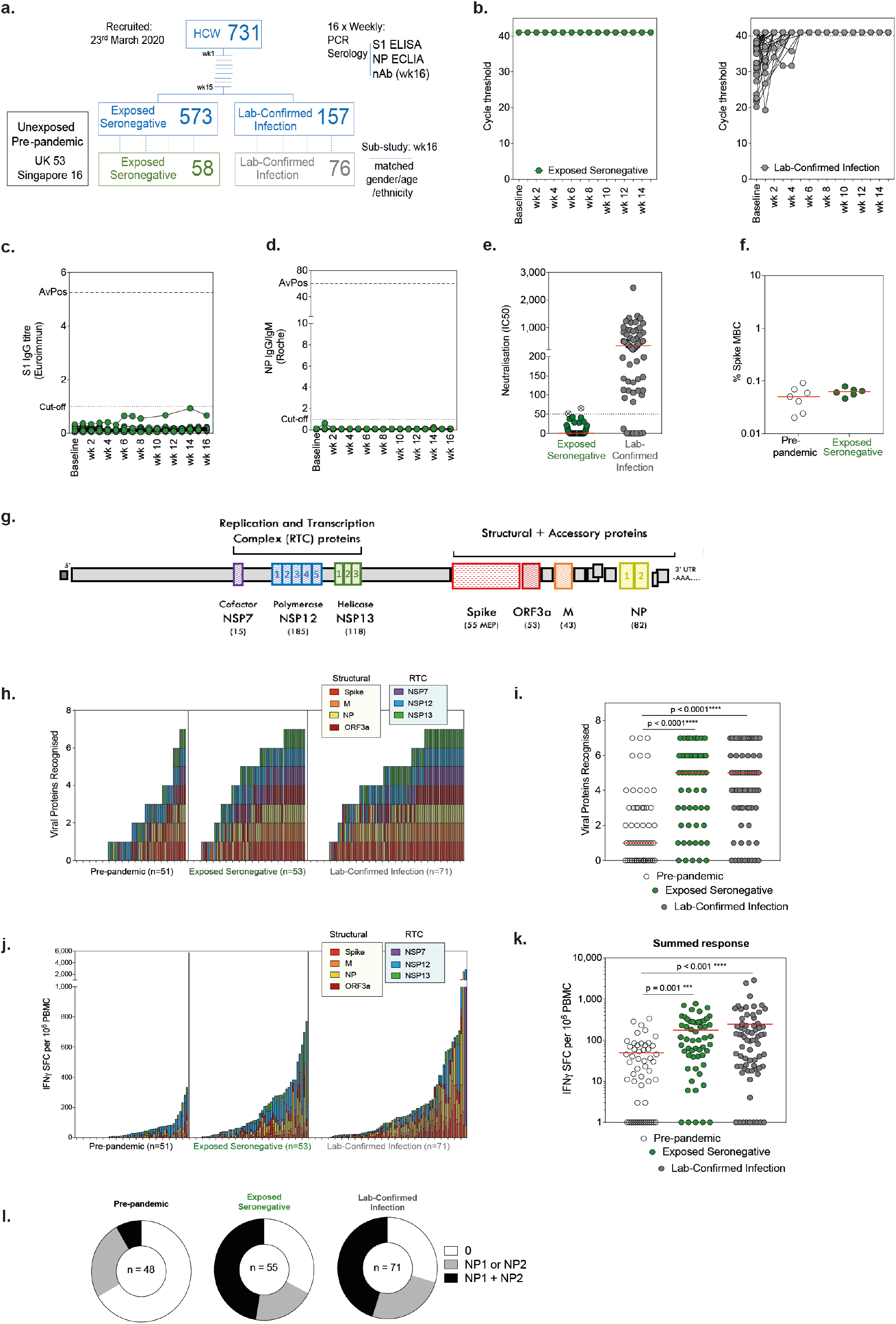
SARS-CoV-2-specific immunity in exposed seronegative HCW. **a**, Design of COVIDsortium prospective HCW study and pre-pandemic cohort. **b**, Longitudinal cycle threshold values for E gene PCR in ES (n=58) and laboratory-confirmed infection (n=76) groups (undetectable at 40 cycles assigned value 41). **c**, Longitudinal anti-Spike S1 and **d**, anti-NP antibody titres in ES (baseline to wk16; n=58; dotted lines at assay positivity cut-off and at average peak [AvPos] response in laboratory-confirmed infected group). **e**, Pseudovirus neutralisation at wk16 (n=58). Crossed circles excluded from ES group (IC50 >50). f, Frequency of SARS-CoV-2 spike-specific memory B cells in pre-pandemic or exposed seronegative cohort (wk16; as a percentage of total memory B cells). **g**, SARS-CoV-2 proteome with RTC and structural regions (and peptide pool numbers) assayed for T cell responses highlighted and number of overlapping 15mer peptides (or mapped epitopes peptides for spike) used in brackets below. **h**, Viral proteins recognised by individuals, coloured by specificity, and **i**, number of viral proteins targeted by group. **j**, Magnitude of T cell response coloured by viral protein and **k**, cumulative magnitude of T cell response by group. Bars, geomean. **l**, Proportion of cohorts with T cell responses to NP1/NP2 pools. **h-l**, IFNγ-ELISpot. **e**,**f**,**i** Bars, median. **i**,**k**, Kruskal-Wallis with Dunn’s correction. ES, exposed seronegative; HCW, health care worker; M, membrane; MEP; mapped epitope pool; NP, nucleoprotein; RTC, replication-transcription complex; SFC, spot forming cells.

We quantified SARS-CoV-2-specific memory T cell responses by ELISpot using the unbiased approach of stimulating PBMC with overlapping peptides covering both structural proteins and the less well-studied key non-structural proteins of the RTC in ORF1ab **(Fig. 1g)**. As previously described, when using sensitive assays^7–9,11,22–25^ (such as IFN -ELISpot with 400,000 PBMC/well used here, some SARS-CoV-2-reactive T cells were detectable in the pre-pandemic samples; however, their multispecificity was significantly lower than in the wk16 laboratory-confirmed infected samples **(Fig. 1h-i;** structural responses at wk16 previously reported^21^). By contrast, ES had SARS-CoV-2-specific T cell responses that were comparable in breadth to the infected HCW at wk16 and significantly more multispecific than in individuals sampled prior to the pandemic **(Fig. 1h-i)**. Not only did ES target more protein pools than pre-pandemics, but they also had an ∼5-fold higher cumulative magnitude of responses than pre-pandemics with an overall strength equivalent to the infected cohort at wk16 **(Fig. 1j-k)**.

We noted that T cells from pre-pandemic samples tended not to target both halves of the NP protein (stimulation pools NP1 & NP2), whereas around 50% of ES and laboratory-confirmed donors targeted both NP pools, confirming our early suggestion^10^that this serves as a simple proxy-measure of a more multispecific response **(Fig. 1l, Extended Data Fig. 1c-d)**. Taken together, we found a higher magnitude and breadth of SARS-CoV-2-specific T cells in repeatedly PCR and antibody negative HCW than in a pre-pandemic cohort.

### RTC-specific T cell and IFI27 signature in ES

Having established that T cell reactivity in the ES group differed from pre-pandemic samples, we next sought to further differentiate them from the group with infection confirmed by PCR and/or seroconversion. Anti-viral T cells recognising immunodominant MHC class I restricted peptides from Flu, Epstein-Barr virus (EBV) and cytomegalovirus (CMV) (FEC) were equivalent between the three cohorts **(Extended Data Fig. 2a)**. However, looking at specificity for structural versus non-structural regions (RTC proteins from ORF1ab) amongst wk16 memory T cells, we found that the relative immunodominance of these regions differed between groups. The infected group had memory T cells dominated by more responses to structural proteins (Spike, membrane, NP, and ORF3a) than to RTC (NSP7, NSP12, NSP13) **(Fig. 2a-b)**. Memory T cells against structural proteins tended to be stronger in those who had a higher viral load, whereas RTC responses did not show this association **(Extended Data Fig. 2b)**. By contrast, pre-existing T cell responses predominantly targeted RTC proteins, whilst ES recognised both regions **(Fig. 2b, Extended Data Fig. 2c-d)**, but with a significantly higher magnitude response to RTC-specific than the infected group **(Fig. 2a, Extended Data Fig. 2d)**. A further small group (11%) of HCW had PCR-confirmed infection but lacked detectable nAb at wk16, some of whom also lacked binding antibodies; interestingly this sub-group was similarly enriched for RTC-reactive T cells **(Extended Data Fig. 2e-f)**.

**Fig. 2.**
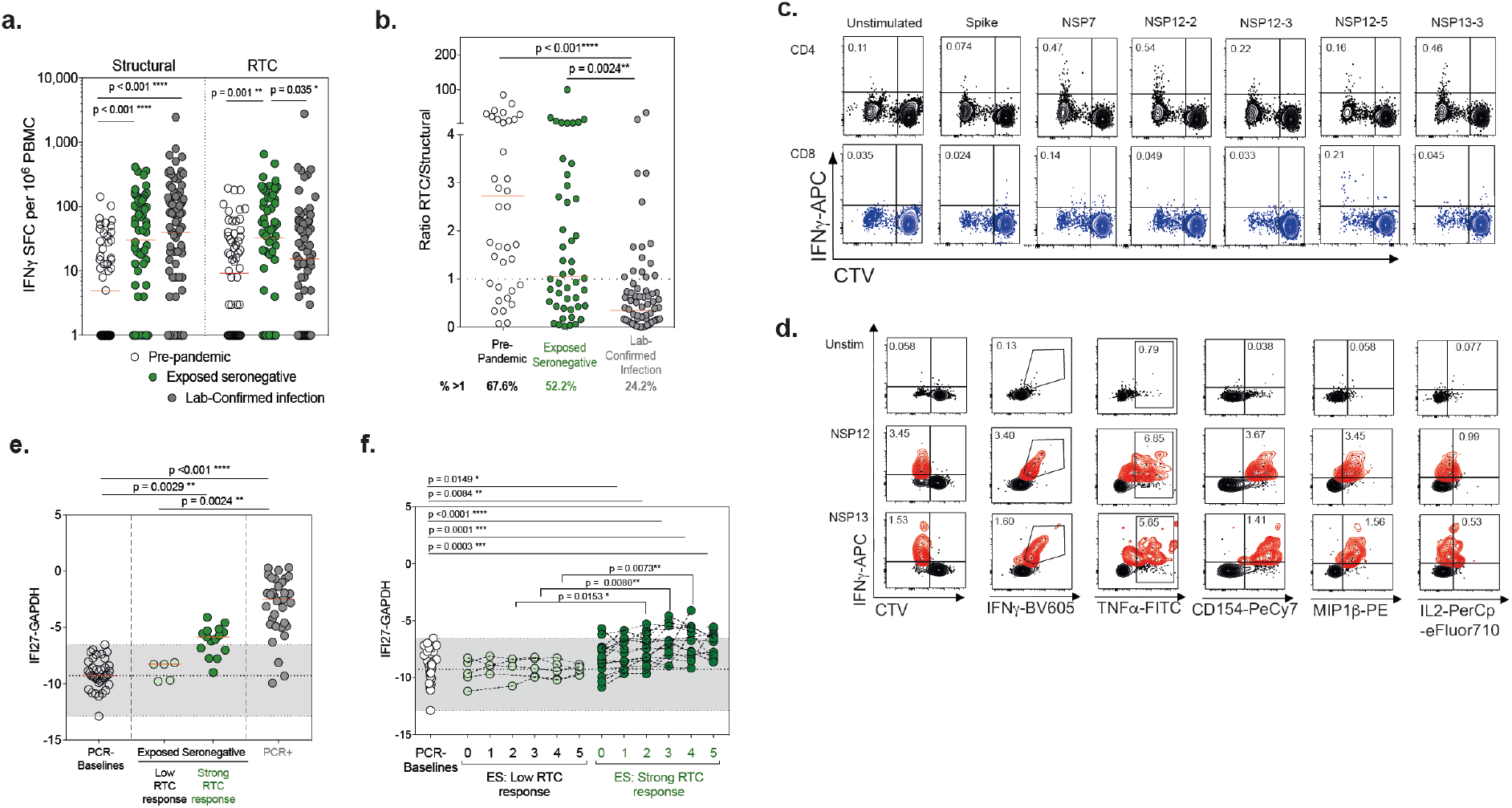
RTC-specific T cell and IFI27 signature in ES: **a**, Magnitude of the T cell response to structural regions and RTC. **b**, Ratio of the T cell response to RTC versus Structural regions. Percentage of cohort with a ratio above 1 (RTC>Structural) shown below. **c**, example CTV and IFN***γ*** staining (gated on CD4+ [black] or CD8+ [blue] T cells) and **d**, dual cytokine or activation marker staining of SARS-CoV-2-specific T cells in an ES after 10-day expansion (proliferating T cells become CTV^lo^ as they divide and dilute out marker) with peptide pools. SARS-CoV-2-specific highlighted in red (CTV^lo^IFN***γ***^+^). Percentage of CD4+ or CD8+ CTV^lo^IFN***γ***^+^ shown. **e**, Peak and **f**, longitudinal (first 5 weeks of follow-up) IFI27 transcript signal by RT-PCR in ES with low (in bottom 20 responders) or strong (in top 20 responders) RTC-specific T cells, compared with other baseline samples from HCW who remained PCR negative throughout, and with HCW at the time of PCR positivity. Range of baseline values highlighted in grey. **a**,**b**, IFNγ-ELISpot. **a**,**b**, bars at geomean. **e**, bars at median. **a**,**b**, Kruskal-Wallis ANOVA with Dunn’s correction.

Taken together, this suggests that the structural proteins, produced in abundance via subgenomic RNA during active infection, are the main targets for T cell responses after mild infection, but that T cells generated by early, transient viral exposure are preferentially focused on the key components of the RTC.

To confirm the T cell identity of ELISpot responses detected in the ES group we expanded them with cognate peptides from RTC regions and used proliferation and cytokine production for flow cytometric identification of antigen-specific CD4+ and CD8+ T cells. T cell responses that divided (CTV dilution) and produced peptide-specific IFNγ could be readily expanded from samples taken from ES at wk16 **(Fig. 2c;** Gating **Extended Data Fig. 3a; Extended data Table 2**). Their post-expansion frequencies tended to be lower than control flu/EBV/CMV-specific responses in the same donors but were in proportion to their differing *ex vivo* frequencies, indicating comparable proliferative potential **(Extended Data Fig. 3b)**. *In vitro* expanded T cell responses in ES were also highly functional, producing multiple cytokines in tandem **(Fig. 2d)**. Most of the SARS-CoV-2-specific T cells expanded from ES were CD4+, however, CD8+ T cell responses were also detectable in most individuals **(Extended Data Fig. 3c)**.

**Fig. 3.**
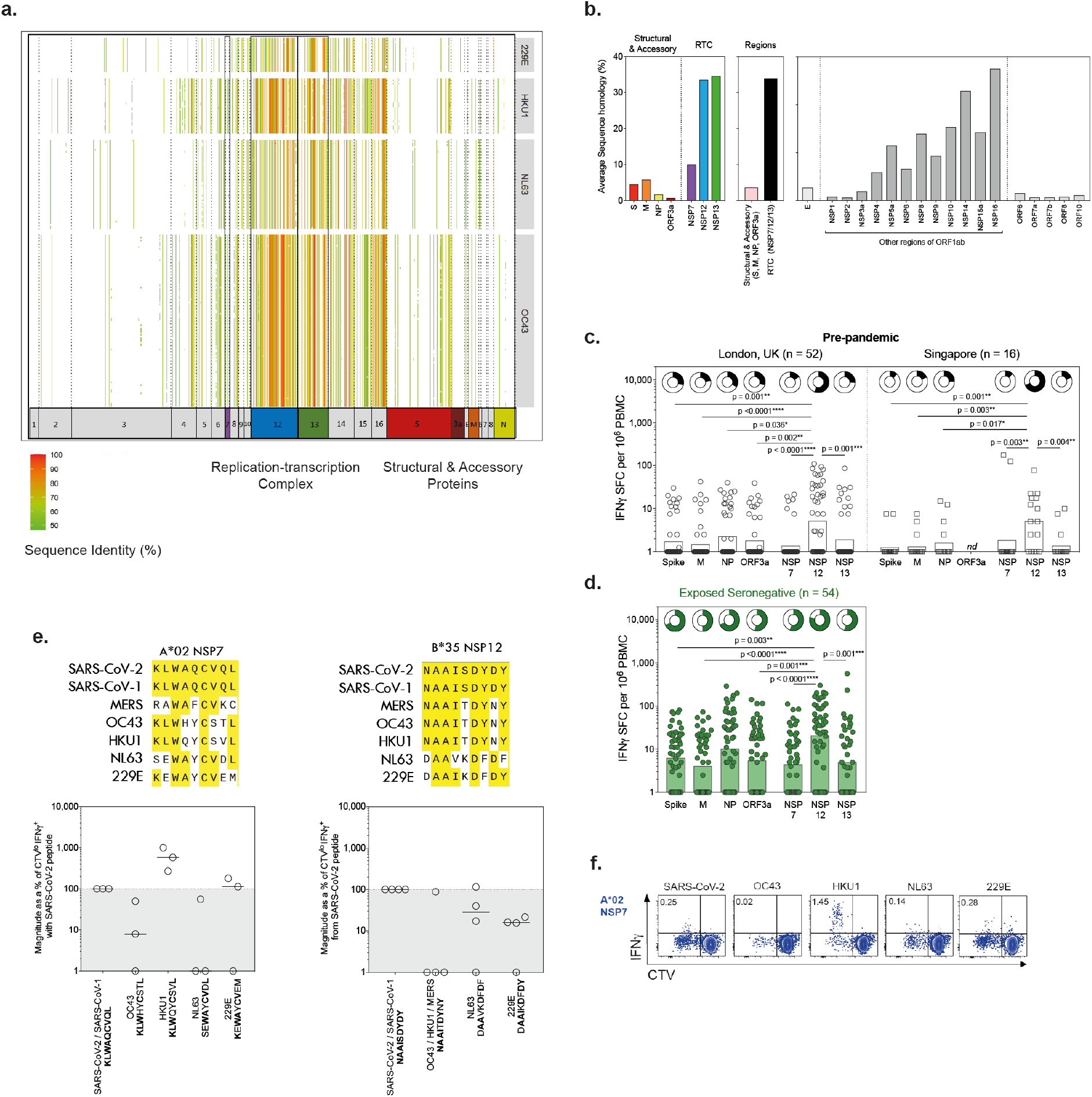
Cross-reactive T cells targeting conserved polymerase: **a**, Heatmap visualising the sequence homology of SARS-CoV-2-derived 15mer peptide sequences to HCoV sequences. Each column represents a 15mer SARS-CoV-2-derived peptide, while each row represents a HCoV genome record. Each cell is coloured by the level of homology of the 15mer peptide to a particular HCoV proteome. Heatmap cells with no fill indicate that no sequence homology greater than 40% was observed. **b**, Average sequence homology of 15mers (overlapping by 14) covering SARS-CoV-2 proteins, or regions (pink, structural [S, M, NP, ORF3a]; black, RTC [NSP7, NSP12, NSP13]), to a set of HCoV sequences (**Supplementary Table 1**). Viral proteins not assayed for T cell responses are shown in grey. **c**, Magnitude of T cell responses to individual SARS-CoV-2 proteins in pre-pandemic samples taken in London, UK and in Singapore and **d**, ES at wk16. Frequency of responders shown in doughnuts above. **e**, Upper panels: alignment of *Coronaviridae* sequences across two CD8 epitopes; conserved amino acids in yellow. Lower panels: Magnitude of CD8+ T cell response (CTV-IFN +) after 10-day expansion with HCoV variant sequence peptides as a percentage of response with SARS-CoV-2 9-mer peptide. **f**, Example plot of CTV vs. IFN -APC after 10-day expansion with SARS-CoV-2 or HCoV sequence 9-mer peptides (ES wk16 samples; gated on single, live, CD3+, CD8+). **c**,**d** Bar, geomean. **e** Bar, median. nd, not done. **c**,**d** Kruskal-Wallis with Dunn’s correction.

Our T cell data raised the possibility that SARS-CoV-2 infection in HCW represents a spectrum, with some ES expanding T cell responses having had a sub-clinical abortive infection not detectable by PCR or antibody seroconversion. To test this postulate, we applied blood transcript measurements of the interferon-inducible gene IFI27 as a biomarker, which we recently showed discriminates early SARS-CoV-2 infection at, or one week before, PCR positivity (specificity 0.95 and sensitivity 0.84^16^). ES with the highest post-exposure RTC-specific responses had significantly raised peak IFI27 levels when compared to baseline controls **(Fig. 2e)**, although levels tended to be lower than in the laboratory-confirmed infected group **(Fig. 2e)**. A time-course of IFI27 over the first five weeks after recruitment showed a stepwise increase in IFI27 in ES, reaching a plateau by week 3-4, by which time almost all first wave laboratory-confirmed infections had occurred **(Fig. 2f)**. By contrast IFI27 was unchanged over 5 weeks sampling in ES with low or undetectable RTC-specific T cell responses **(Fig. 2f)**. Therefore, a low-level systemic interferon response indicative of virus exposure was detectable in individuals who had the strongest SARS-CoV-2-specific T cell response post-exposure, despite them lacking PCR or antibody confirmation of SARS-CoV-2 infection. Extrapolating from our previous data showing that IFI27 is induced at the time of incident infection and correlates with viral load^16^, this supports the ES with stronger RTC-specific T cell responses representing HCW who have experienced a low-level/transient infection. Thus, ES with SARS-CoV-2 T cell reactivity could be distinguished by both their innate IFI27 signature and the propensity of their T cells to target RTC.

### Cross-reactive T cells targeting conserved polymerase

A transient/abortive infection not detectable by PCR or seroconversion could conceivably result from a lower viral inoculum and/or from a more efficient innate and/or adaptive immune response. The latter would be favoured by pre-existing memory T cell responses with the potential to expand rapidly upon cross-recognition of early viral products of SARS-CoV-2 replication. Early T cell proliferation and TCR clonal expansion, even prior to detectable virus, has been observed during mild SARS-CoV-2^22,38^ and expansion of virus-specific T cells predates antibody induction after mRNA vaccination^39^. Having found the ES group to be enriched for SARS-CoV-2-specific T cells, particularly against RTC, we therefore investigated the possibility that some of these represented expansions of pre-existing cross-reactive responses.

Likely candidates for the induction of T cells that cross-recognise SARS-CoV-2 are closely related human endemic common cold coronaviruses (HCoV: α-HCoV-229E and NL63, and β-HCoV HKU1 and OC43). We bioinformatically determined the sequence homology of all possible SARS-CoV-2-derived 15mer peptides to a curated set of HCoV sequences (**Supplementary Table 1**). We found that the RTC proteins, expressed at the first stage of the SARS-CoV-2 life cycle^15^, have 15mer sequences that are of high homology to the HCoVs **(Fig. 3a**)^24,40^. In particular, NSP7, NSP12, and NSP13-derived 15mers had 6.3, 29.9 and 31.0% higher average sequence homology to the four HCoVs compared to structural protein-derived 15mers (all p<0.001, **Fig. 3b**). NSP12 being the largest of these 3 proteins, represents the region with the most homology overall. Interestingly, the highly conserved RNA polymerase (NSP12) was also the region that was found to be most commonly targeted when screening our cohort of 52 pre-pandemic samples. T cell responses to NSP12 showed the highest average magnitude and frequency of donors responding **(Fig. 3c)**. Of note, the same preferential targeting of NSP12 was observed in a geographically distinct cohort of pre-pandemic samples from Singapore **(Fig. 3c)**.

Pre-existing T cells had the potential to recognise all viral antigens tested, including those with less conservation across HCoV, as previously described^22,41^ and responses against all these regions were further enriched in ES, suggesting many sources of pre-existing responses and *de novo* generated responses can contribute to T cell memory in exposed seronegative individuals. However, as with pre-pandemics samples, ES preferentially targeted NSP12 **(Fig. 3d)**. Therefore, the viral protein most commonly targeted by pre-existing T cells is also the largest conserved region, suggesting that exposure to HCoV is likely one source of cross-reactive T cells.

To further explore the potential for cross-reactivity due to prior infection with seasonal HCoV in this group, we first carried out epitope mapping of RTC-specific T cells from ES. Two-dimensional mapping matrices were used to determine individual immunogenic 15mers (Example plots **Extended Data Fig. 3d; Supplementary Table 2**), which identified novel and previously described epitopes post-infection or in pre-pandemic samples^8,10,25,42^(**Extended Data Table 3**). Next, we aligned viral sequences for HCoV at the CD4+ and CD8+ epitopes we had identified in ES, which highlighted sequence conservation at the level of the individual T cell epitope (CD4+: **Extended Data Fig. 3e;** CD8+: **Fig. 3e**).

We selected individuals from the ES cohort with the relevant HLA type to test whether their CD8+ T cells could respond to the seasonal HCoV variant peptide sequences from the RNA polymerase and RNA-polymerase cofactor epitopes. We identified a clear example of a cross-reactive T cell response against the HLA-A*02:01 restricted epitope in RNA-polymerase cofactor NSP7; T cells from all three HLA-A*02:01+ ES HCW tested had stronger responses to the HKU1 sequence (KLWQYCSVL) than to seasonal HCoV or SARS-CoV-2 **(Fig. 3e;** Example plots **Fig. 3f)**. This suggested prior HKU1 infection primed these NSP7 responses, which were then able to cross-recognise the SARS-CoV-2 sequence, albeit with reduced efficiency. All four HLA*B35 ES also showed some cross-recognition of seasonal HCoV variant epitopes in NSP12, with the extent varying as would be expected in light of heterogeneity in previous HCoV exposure and T cell repertoire composition **(Fig. 3e)**.

In summary, key RTC regions like the RNA polymerase, that are expressed in the first stage of the viral life cycle, are highly conserved among HCoV and are preferentially targeted by T cells in pre-pandemic and ES samples. T cells from donors able to abort infection can cross-recognise SARS-CoV-2 and HCoV sequences at individual epitopes within the RTC, pointing to prior infection with HCoV as a source of some pre-existing cross-protective T cells.

### Preferential expansion of polymerase-specific T cells in abortive infection

To examine whether pre-existing cross-reactive and/or rapidly generated *de novo* RTC-specific T cells can expand *in vivo*, we took advantage of unusual access to paired PBMC samples taken pre-and post-SARS-CoV-2 exposure. Firstly, we recruited a cohort of medical students and laboratory staff from whom stored PBMC were available from winter 2018-2019 (n = 23), prior to the COVID-19 pandemic, and sampled them again after known close contact with infected cases, with or without IgG seroconversion +/-PCR positivity (**Extended Data Table 4**). Pre-and post-exposure/infection PBMC were analysed in parallel for ELISpot responses to RTC and structural pools.

There was clear evidence for *in vivo* expansion of pre-existing NSP12 responses in 4/5 individuals who had exposure to SARS-CoV-2 through a close contact but who remained seronegative, with three cases showing a more than two-fold expansion **(Fig. 4a-b)**. The other five seronegative close contacts had no pre-existing NSP12 responses detectable, but four of these had detectable, presumed *de novo*, low-level responses after exposure **(Fig. 4b)**. Overall, the close-contact seronegative group showed preferential expansion of RTC over structural protein responses comparing their pre-and post-exposure samples **(Fig. 4a-b)**. By contrast, the group with serological confirmation of infection also showed the expected *in vivo* expansion of pre-existing SARS-CoV-2-reactive T cells but this was predominantly of responses directed against structural proteins. Only four out of thirteen in the seroconverted group expanded NSP12 responses and overall, they had no significant increase in RTC-specific T cells **(Fig. 4a; Extended Data Fig. 4a**).

**Fig. 4.**
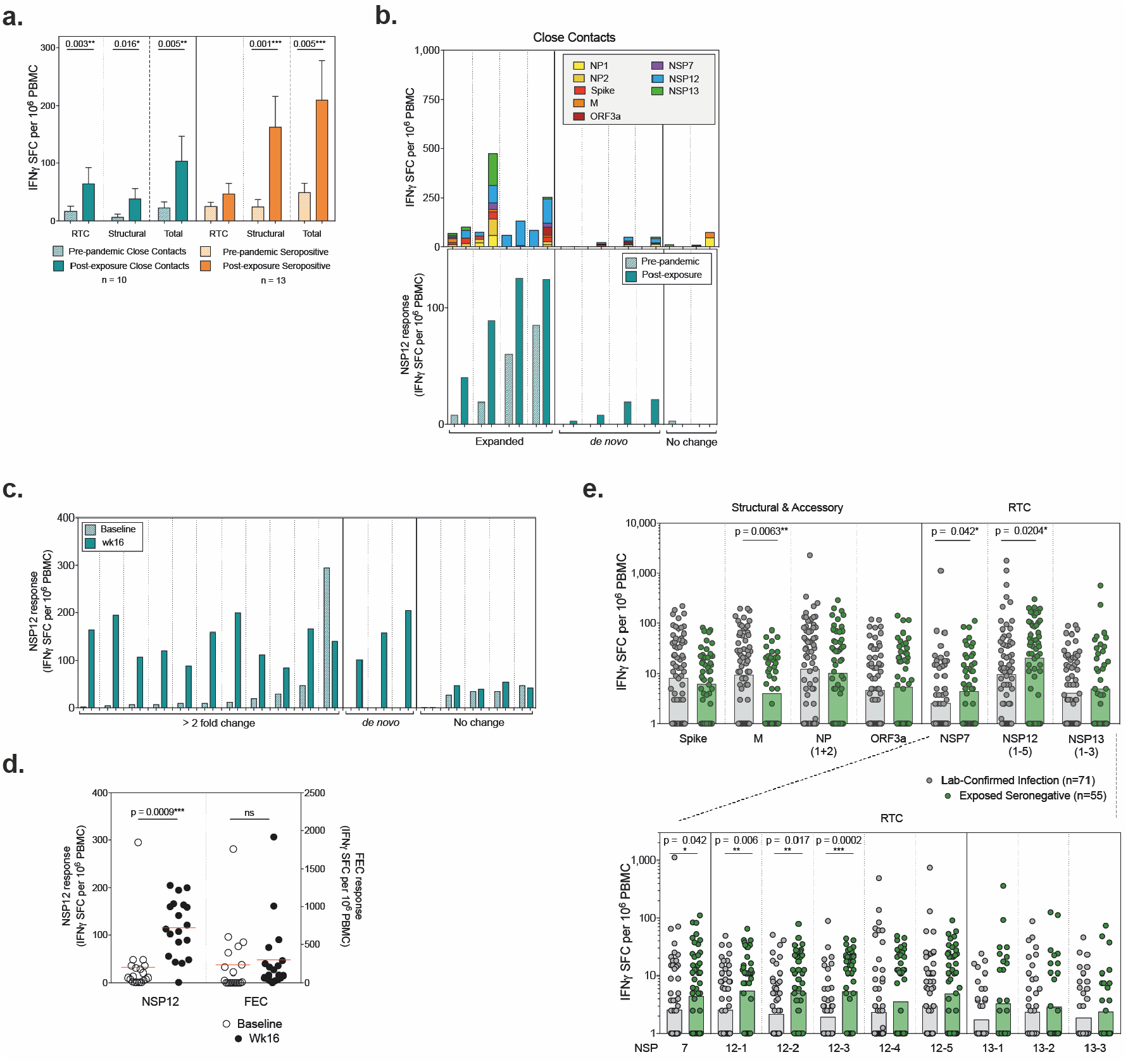
*In vivo* expansion of polymerase-specific T cells in abortive infection: **a**, Magnitude of T cell response to RTC, structural proteins, and total response in seronegative close contacts of cases, or in seropositive infected individuals. Bar, mean + SEM. **b**, Change in magnitude of T cell response between pre-pandemic and post-exposure samples (upper panel: all proteins, lower panel: NSP12) from seronegative close contacts of cases, coloured by protein target. **c**, Change in magnitude of NSP12 T cell response between recruitment and post-exposure (wk16) in ES (sub-group with the top 19 RTC response at wk16). **d**, Change in magnitude of NSP12 and Flu, EBV, CMV (FEC) responses between recruitment and wk16 in ES (sub-group with the top 19 RTC response at wk16). Bars, mean. Wilcoxon t-test. **e**, Magnitude of T cell response to all individual SARS-CoV-2 proteins tested (upper panel) and to sub-pools of equivalent length (∼40 overlapping peptides; lower panel) within RTC at wk16 in laboratory-confirmed infected HCW or ES. Bars, geomean. Mann-Whitney test.

We then reverted to the ES HCW cohort, where small volume PBMC collections were available from the time of recruitment, allowing targeted analysis of baseline responses in those with the strongest RTC responses at wk16.

Due to rapid recruitment HCW sampling started the week infections peaked in London (UK, 23^rd^ March 2020), meaning most SARS-CoV-2 exposure was around the time of recruitment (79% of positive PCR tests within first 2 weeks of follow-up, no PCR+ after week 5 of follow-up, **Fig. 1b**)^16,43^, with seroconversion within the first 3 weeks of follow-up for most^36^. Focusing on the most common specificity of pre-existing responses, NSP12-specific T cells were already detectable at baseline in 74% of those ES with the strongest NSP12 responses post-exposure **(Fig. 4c)**. NSP12 responses expanded *in vivo* on average 8.4-fold between recruitment and wk16 of follow-up, with no corresponding change in Flu/EBV/CMV responses **(Fig. 4d)**. We could identify three ES with *de novo* responses, ten with >2 fold-expansion of NSP12-specific T cells and one with a >2 fold-contraction, in line with their reported likely exposure before recruitment **(Fig. 4c)**.

Interestingly, all HCW with *de novo* or expanded/contracting NSP12-specific T cell responses also had NP1 and NP2 reactive T cells after exposure **(Extended Data Fig. 4b)**; however, of the five individuals who had no change in NSP12 response only 2/5 had these specificities, suggesting they may not have had the same level of SARS-CoV-2 exposure. The fold-change in NSP12 between recruitment and wk16 follow-up correlates with the total SARS-CoV-2 response, suggesting it may be used as a proxy to identify exposed seronegative individuals who have had expanded T cell immunity after exposure **(Extended Data Fig. 4c)**.

Finally, we compared the magnitude of memory T cell responses of different specificities at wk16 to see if there was a preferential enrichment of RTC-specific responses in ES HCW compared to the laboratory-confirmed infected HCW. Strikingly, NSP12 was the only viral protein to induce T cells to a higher magnitude in the cohort of seronegative individuals in which a successful infection was not established compared to those with classical laboratory-confirmed infection **(Fig. 4e)**. Examining the RTC specificities in more detail (breaking down according to peptide pools of equivalent size, ∼40 overlapping 15mers) revealed that T cell responses targeting several regions of the RNA polymerase NSP12 were significantly enriched in the exposed seronegative HCW cohort compared to post-infection, whilst other RTC pools also showed non-significant trends for enrichment **(Fig. 4e**, lower panel) suggesting that they may have played a role in protecting these individuals from PCR-detectable infection and seroconversion.

In summary, we provide T cell and innate transcript evidence for abortive, seronegative SARS-CoV-2 infection. Pre-existing cross-reactive T cells, in particular against the RNA-polymerase, expanded *in vivo* following SARS-CoV-2 exposure, and were preferentially enriched in individuals in whom SARS-CoV-2 failed to establish a successful infection, compared to those with classical infection. Taken together, these data highlight a role for the *in vivo* expansion of pre-existing and *de novo* RTC-specific T cells in aborting early viral infection before the induction of antibodies.

## Conclusions

The first proteins to be transcribed during SARS-CoV-2 replication are those of the RTC within ORF1ab^13^, which may make them effective targets for early viral control. Non-structural proteins of ORF1ab are released into the cytoplasm as part of the viral life cycle^13^, giving direct access to the major histocompatibility complex class (MHC) I presentation pathway to activate CD8+ T cell responses to these regions, as has been shown for other single strand RNA viruses including dengue^30^. The formation of the RTC is essential for subsequent transcription of the viral genome, raising the possibility that some infected cells could be recognised and removed by CD8+ T cells before widespread production of structural proteins and mature virion formation^44^. Whereas live virus is likely to be more effectively presented in MHC class I, exogenous viral antigen, for instance from non-replicative particles, can lead to the priming or activation of CD4+ T cells.

The differential biasing of T cells towards early expressed viral proteins at the expense of humoral responses and T cells targeting structural proteins in HCW not seroconverting may reflect repetitive occupational exposure to very low viral inocula, as has been reported in HIV and SIV^27,32,45^. Consistent with this hypothesis, we did note some *de novo* induction of T cells not detectable prior to exposure in the ES. However, we also documented expansion of pre-existing T cells against the highly conserved polymerase, with responses capable of cross-recognising epitope variants between seasonal HCoV and SARS-CoV-2. Such pre-existing T cells, at higher frequency than naïve T cells and poised for immediate re-activation on encountering their antigen, would be expected to favour abortive infection. HCW are particularly prone to exposure to respiratory pathogens^46–48^ and have higher frequencies of HCoV-reactive T cells than the general public^19^. Recent HCoV infection is associated with reduced risk of severe COVID-19 infection^49^, likely partly attributable to cross-reactive neutralising antibodies^50,51^, however, pre-existing T cell responses have also been suggested to reduce the risk of subsequent infection^52^. Supporting a role for *de novo* or pre-existing T cell responses in early control of SARS-COV-2, we have recently shown that a T cell proliferation signature can be detect prior to PCR positivity in those with mild COVID-19 and this is accompanied by rapid expansion of SARS-CoV-2-specific TCRs^38^.

CD4+ T cell responses are more likely to cross-recognise similar viral sequences due to greater flexibility in peptide binding within MHC Class II^53^, in line with the dominance of CD4 responses in ES shown here. However, cross-reactive CD8+ T cells are also well-described, with Nelde *et al* estimating that 70% of epitopes recognised by pre-existing T cell responses had physico-chemical similarities to HCoV sequences and approximately 50% of the epitopes identified by Ferretti *et al* lying within ORF1ab in areas of low mutational variation^8,25^. Essential viral proteins, such as the RTC, that have less scope to mutate whilst retaining functionality, are more conserved across the *Coronaviridae* family than structural regions^40^ and therefore retain T cell epitopes. Preferential involvement of pre-existing responses, dominated by RTC-specific T cells, in early control would explain their enrichment after abortive infection compared to classical infection.

Although we have shown an association between the presence of certain T cell specificities, in particular to areas of high conservation within HCoV such as NSP12, and resistance to overt infection in exposed HCW, larger cohorts or human challenge studies will be needed to determine their relative contribution to protection. The antiviral potential of CD8+ T cells in SARS-CoV-2 is supported by depletion experiments in macaques^54^ and by the resolution of infection in patients lacking humoral immunity because of agammaglobulinemia or B cell depletion therapy^55,56^. It remains possible that innate control alone can mediate abortive infection, with low level antigen production being enough to generate RTC-biased T cell responses simply as a biomarker of low-grade infection. The fact that the ES cohort had much lower levels of IFI27, reflective of less interferon induction, than the infected cohort does not support preferential innate control in the former. However, interferon-independent induction of RIG-I has been proposed as another potential mechanism of aborting SARS-CoV-2 infection^15^. A further caveat in the interpretation of our findings is that we only analysed peripheral immunity; it is plausible that mucosal-sequestered antibodies, recently reported in seronegative HCW^57^, could have played a role in our seronegative cohort.

We have described an under-investigated host-pathogen interaction leading to the induction of innate and cellular immunity without seroconversion. These data will inform the design of studies where true unexposed comparator groups are required and highlight a key population of individuals where risk of SARS-CoV-2 reinfection and immunogenicity of vaccines should be independently assessed. Our data suggest T cells recognising the RTC are particularly effective at early control of infection and may offer pan-*Coronaviridae* reactivity against both endemic and emerging viruses, arguing for their inclusion and assessment in next-generation vaccines.

## Data Availability

All relevant data analysed during this study are included in this published article (and its supplementary information files). Custom scripts used to perform the homology searches, heatmap visualisation and permutation testing are hosted on GitHub (https://github.com/cednotsed/tcell_cross_reactivity_covid.git). The datasets generated during and/or analysed during the current study are available from the corresponding author on reasonable request. Correspondence and requests for materials should be addressed to MKM or LS.

## Methods

### COVIDsortium Healthcare Worker Participants

The COVIDsortium bioresource was approved by the ethical committee of UK National Research Ethics Service (20/SC/0149) and registered on ClinicalTrials.gov (NCT04318314). Full study details of the bioresource (participant screening, study design, sample collection, and sample processing) have been previously described^21,58^.

In this cohort and London as a whole infections peaked during the first week of lockdown (March 23^rd^ 2020)^43^, and we observed approximately synchronous exposure coincident with recruitment, we therefore used this as the benchmark for assessing exposure generated immunity. Across the main study cohort, 48 participants had positive RT-PCR results with 157 (21.5%) seropositive participants. Infections were asymptomatic or mild with only two hospital admissions (none requiring intensive care admission). The cross-sectional case controlled sub-study (n=129) collected samples at 16-18 weeks after the first UK lockdown **(Fig. 1a)**. Power calculations were performed prior to week 16 sub-study sampling to determine the sample size needed to test the hypothesis that HCW with pre-existing T cell responses are enriched in exposed uninfected group at a range of incidence of infection, assuming 50% of cohort had pre-existing T cell responses. Sample sizes of 18-64 per group were estimated. An age, sex and ethnicity matched nested sub study was designed within the larger (n=731) parent study and 129 attended for 16-week sampling including high volume PBMC isolation.

Lab confirmed infection was determined by weekly nasopharyngeal RNA stabilizing swabs and reverse transcriptase polymerase chain reaction (RT-PCR; Roche cobas SARS-CoV-2 test, Envelope [E] gene) and antibody assay positivity (Spike protein 1 IgG Ab assay, EUROIMMUN) and anti-nucleocapsid total antibody assay (ROCHE) described in detail below. The exposed seronegative health care worker cohort were matched for demographics and exposure to the lab confirmed infected cohort and was defined by negativity by these three tests at all 16 time points as well as negative for neutralising antibodies at week 16 and at selected prior time points as indicated.

The cohort of medical students and laboratory staff was approved by UCL Ethics (Project ID Number: 13545/001) and pre-pandemic healthy donor samples were collected and cryopreserved before August 2019 under ethics numbers 11/LO/0421. All subjects gave written informed consent and the study conformed to the principles of the Helsinki Declaration.

### Isolation of PBMC and Serum

Peripheral blood mononuclear cells (PBMC) were isolated from heparinized blood samples using Pancoll (Pan Biotech) or Histopaque®-1077 Hybri-MaxTM (Sigma-Aldrich) density gradient centrifugation in SepMate tubes (StemCell) according to the manufacturer’s specifications. Isolated PBMCs were cryopreserved in fetal calf serum containing 10% DMSO and stored in liquid nitrogen.

Whole blood samples were collected in SST vacutainers (VACUETTE) with inert polymer gel for serum separation and clot activator coating. After centrifugation at 1000 X g for 10 min at room temperature (RT), serum layer was aliquoted and stored at -80 °C. All T cell assays reported here were performed on cryopreserved PBMC.

### Weekly SARS-CoV-2 S1 and NP Serology

Weekly Euroimmun anti-SARS-CoV-2 enzyme-linked immunosorbent assay (ELISA; anti-SARS-CoV-2 S1 antigen IgG and the Roche Elecsys anti-SARS-CoV-2 electrochemiluminescence immunoassay (ECLIA; anti-SARS-CoV-2 nucleoprotein IgG/IgM) commercial assays were performed by Public Health England as previously described^21^. S1 ELISA: A ratio of ≥ 1.1 was deemed positive. A ratio of 11 was taken to be the upper threshold as the assay saturates beyond this point. NP ECLIA: Anti-NP results are expressed as a cut-off index (COI) value based on the electrochemiluminescence signal of a two-point calibration, with results COI ≥1.0 classified as positive.

### Neutralization assays – Pseudotype and authentic virus

SARS-CoV-2 pseudotype neutralisation assays were conducted using pseudotyped lentiviral particles as previously described^21^. Briefly, serum was heat-inactivated at 56 °C for 30 mins. Serum dilutions in DMEM were performed in duplicate with a starting dilution of 1 in 20 and 7 consecutive 2-fold dilutions to a final dilution of 1/2,560 in a total volume of 100 µl. 1 × 10^5^ RLU of SARS-CoV-2 pseudotyped lentiviral particles were added to each well (serum dilutions and controls) and incubated at 37 °C for 1 hr. 4 × 10^4^ Huh7 cells suspended in 100 μl complete media were added per well and incubated for 72 hr at 37 °C and 5% CO_2_. Firefly luciferase activity (luminescence) was measured using Steady-Glo^®^ Luciferase Assay System (Promega) and a CLARIOStar Plate Reader (BMG Labtech). The curves of relative infection rates (in %) versus the serum dilutions (log10 values) against a negative control of pooled sera collected prior to 2016 (Sigma) and a positive neutraliser were plotted using Prism 9 (GraphPad). A non-linear regression method was used to determine the dilution fold that neutralised 50% (IC50).

Authentic SARS-CoV-2 microneutralization assays were carried out as previously described^59^. Briefly, a mixture of serum dilutions in DMEM (1 in 20 and 11 consecutive 2-fold dilutions to a final dilution of 1/40,960) and 3 × 10^4^ FFU of SARS-CoV-2 virus (Wuhan Hu-1) were incubated at 37 °C for 1 hr. After initial incubation, pre-seeded Vero E6 cells were infected with the serum-virus samples and incubated (37 °C and 5% CO_2_) for 72 hr. Cells were then fixed with 100 μl 3.7% (vol/vol) formaldehyde for 1 hr. Cells were washed with PBS and stained with 0.1% (wt/vol) crystal violet solution for 10 minutes. After removal of excess crystal violet and air drying, the crystal violet stain was re-solubilized with 100 μl 1% (wt/vol) sodium dodecyl sulfate solution. Absorbance readings were taken at 570 nm using a CLARIOStar Plate Reader (BMG Labtech). Absorbance readings for each well were standardised against technical positive (virus control) and negative (cells only) controls on each plate to determine a percentage neutralisation value. A non-linear regression (curve fit) method was used to determine the dilution fold that neutralised 50% (IC50) using Prism 9 (GraphPad). SARS-CoV-is classified as a hazard group 3 pathogen and therefore all authentic SARS-CoV-2 propagation and microneutralization assays were performed in a containment level 3 facility.

### Spike ELISA

Seropositivity against SARS-CoV-2 spike was determined for medical student and laboratory staff cohort between July 2020 and Jan 2021 (**Extended Data Table 4**) by enzyme-linked immunosorbent assay, as validated and described previously^50,60,61^. Briefly, 9 columns of 96-half-well MaxiSorp plates (Thermo Fisher Scientific) were coated overnight at 4 °C with purified S1 protein in PBS (3 μg/ml per well in 25 μl), the remaining 3 columns were coated with goat anti-human F(ab)’2 (1:1,000) to generate in internal standard curve. The next day, plates were washed with PBS-T (0.05% Tween in PBS) and blocked for 1 hr at RT with assay buffer (5% milk powder PBS-T). Sera were diluted in blocking buffer (1:50). 25 ul of serum was then added to S1 coated wells in duplicate and incubated for 2 hr at RT. Serial dilutions of known concentrations of IgG were added to the F(ab)’2 IgG-coated wells in triplicate (Sigma Aldrich). Following incubation for 2 hr at RT, plates were washed with PBS-T and 25 µl alkaline phosphatase-conjugated goat anti-human IgG (Jackson ImmunoResearch) at a 1:1000 dilution in assay buffer added to each well and incubated for 1 hr RT. Plates were then washed with PBS-T, and 25 µl of alkaline phosphatase substrate (Sigma Aldrich) added. ODs were measured using a MultiskanFC (Thermofisher Scientific) plate reader at 405 nm and **S1**-specific IgG titers interpolated from the IgG standard curve using 4PL regression curve-fitting on GraphPad Prism 8.

### SARS-CoV-2 spike-specific Memory B cell staining

Multiparameter flow cytometry was used for *ex vivo* identification of spike-specific memory B cells staining as previously described^37^. Biotinylated tetrameric spike (1 ug) was fluorochrome linked by incubating with streptavidin conjugated APC (Prozyme) and PE (Prozyme) for 30 mins in the dark on ice. PBMC were thawed and incubated with Live/Dead fixable dead cell stain (UV, ThermoFisher Scientific) and saturating concentrations of phenotyping mAbs diluted in 50% 1 x PBS 50% Brilliant Violet Buffer (BD Biosciences): CD3 Bv510 (Biolegend, clone OKT3), CD11c FITC (BD Biosciences, clone B-ly6), CD14 Bv510 (Biolegend, clone M5E2), CD19 Bv786 (BD bioscience, clone HIB19), CD20 AlexFluor700 (BD biosciences 2H7), CD21 Bv711 (BD biosciences, clone B-ly4), CD27 BUV395 (BD biosciences, clone L128), CD38 Pe-CF594 (BD biosciences, clone HIT2), IgD Pe-Cy7 (BD biosciences, clone IA6-2). For identification of SARS-CoV-2 antigen specific B cells 1 μg per 500 μl of stain each of tetrameric Spike-APC and Spike-PE were added to cells. Cells were incubated in the staining solution for 30 mins RT, washed with PBS, and subsequently fixed with FoxP3 Buffer Set (BD Biosciences) according to the manufacturer’s instructions. All samples were acquired on a BD Fortessa-X20 flow cytometer. Data were analysed by FlowJo version 10.7 (TreeStar). Example gating and positivity cut-off have been previously reported^37^. The magnitude of the SARS-CoV-2 spike-specific memory B cell population is expressed as a percentage of memory B cells (gated as: lymphocytes, singlets, Live, CD3-CD14-CD19+, CD20+, excluding: CD38^hi,^ IgD+ and CD27+CD21-) binding both PE and APC labelled spike.

### SARS-CoV-2 peptides

Full lists of the peptides contained in pools of overlapping peptides covering structural^21^and RTC proteins^10^have been previously described. 15-mer peptides overlapping by 10 amino (GL Biochem Shanghai Ltd, >80% purity). Overlapping peptides of NSP12 are listed in **Supplementary Table 3**. For IFN -ELISPot assays SARS-CoV sequence peptides were used (96.5% sequence homology with Wuhan SARS-CoV-2 consensus sequence, 34/931 amino acids differ). For epitope mapping SARS-CoV-2 sequence peptides were used for NSP12-2 and NSP12-5 (GL Biochem Shanghai Ltd, >80% purity**; Supplementary Table** 3).

To limit competition for *in vitro* for peptide presentation we limit stimulations to a maximum of 55 peptides and have, therefore, divided large proteins such as NP into sub-pools: NP (NP1, NP2, 41 peptides each), M (43 peptides), ORF3a (53 peptides), NSP7 (15), NSP12 (36-37 per pool NSP12-1 to NSP12-5) and NSP13 (39-40 peptides per pool NSP13-1 to NSP13-3). In addition 15-mer peptides covering the predicted SARS-CoV-2 spike epitopes^10^ to give a total of 55 peptides in this pool (Spike). Optimal 9mer peptides for CD8+ epitopes were custom synthesised by ThinkPeptides (UK) >70% purity.

### IFNγ-ELISpot Assay

IFNγ-ELISpot Assay was performed as previously described on cryopreserved PBMC^10,21,62^. Unless otherwise stated, culture medium for human PBMC was sterile 0.22 μM filtered RPMI medium (Thermo Fisher Scientific) supplemented with 10% by volume heat inactivated (1 hr, 64 °C) fetal calf serum (FCS; Hyclone, and 1% by volume 100 x penicillin and streptomycin solution (GibcoBRL).

ELISpot plates (Merck-Millipore, MSIP4510) were coated with human anti-IFNγ Ab (1-D1K, Mabtech; 10 μg/ml) in PBS overnight at 4 °C. Plates were washed 6x with sterile PBS and were blocked with R10 for 2 hr at 37 °C with 5% CO_2_. PBMC were thawed and rested in R10 for 3 hr at 37 °C with 5% CO_2_ before being counted to ensure only viable cells were included. 400,000 PBMC were seeded in R10/well and were stimulated for 16-20 hr with SARS-CoV-2 peptide pools (2 μg/ml/peptide) at 37 °C in a humidified atmosphere with 5% CO_2_. Where insufficient cells were available NSP12 pools 1,2 and 3 and NSP13 pools 1,2,3 were combined into a single well. HCW who did not have a full complement of stimulations were excluded from analysis of total magnitude of breadth of response, hence slightly lower n numbers. Internal plate controls were R10 alone (without cells) and two DMSO wells (negative controls), concanavalin A (ConA, positive control; Sigma-Aldrich) and FEC (HLA I-restricted peptides from influenza, Epstein-Barr virus, and CMV; 1 μg/ml/peptide). ELISpot plates were developed with human biotinylated IFN-γ detection antibody (7-B6-1, Mabtech; 1μg/ml) for hr at RT, followed by incubation with goat anti-biotin alkaline phosphatase (Vector Laboratories; 1:1000) for 2 hr RT, both diluted in PBS with 0.5% BSA by volume (Sigma-Aldrich), and finally with 50 μl/well of sterile filtered BCIP/NBT Phosphatase Substrate (ThermoFisher) for 7 min RT. Plates were washed in ddH_2_0 and left to dry overnight before being read on an AID classic ELISpot plate reader (Autoimmun Diagnostika GMBH, Germany).

The average of two DMSO wells was subtracted from all peptide-stimulated wells for a given PBMC sample and any response that was lower in magnitude than 2 standard deviations of these sample specific DMSO control wells was not considered a peptide specific response (given value 0). Results were expressed as IFNγ spot forming cells (SFC) per 10^6^ PBMC after background subtraction. The geometric mean of all DMSO wells was 9.571 SFC per 10^6^PBMC (3.8 spots). We excluded the results if negative control wells had >95 SFC/10^6^ PBMC or positive control wells (ConA) were negative. T cell responses to SARS-CoV-2 did not correlate with background spots in DMSO wells (e.g. ES cohort spearman r = -0.068 p = 0.6141).

### Antigen-specific T cell proliferation assay and epitope mapping

Frozen PBMC were thawed and washed twice with sterile PBS. PBMC were resuspended in 1 mL R10 culture media (2-10 × 10^6^ PBMC) and 0.5 µL of 5 mM stock CellTrace violet (CTV; Thermo Fisher Scientific) was added per sample with mixing. PBMC were stained in the dark for 10 mins at 37 °C in a humidified atmosphere with 5% CO_2_. Ten-times volume of cold R10 was added to stop the staining reaction, and cells were incubated for 5 mins on ice. Cells were washed in PBS and incubated for 5 mins at 37 °C before being transferred to a new tube and were washed again in R10. CTV stained PBMC were plated in 96-well plates (2-4 × 10^5^ PBMC in 200 µL R10) and stimulated with peptide pools (2 μg/ml per peptide) for 10 days in R10 supplemented with 0.5 μg/ml soluble anti-CD28 (Thermo Fisher scientific) and 20 U/ml recombinant human IL2 (Peprotech). A Small sample of CTV-stained and unstained PBMC were run to confirm efficiency of staining. 100 µL media was removed and replaced with R10 supplemented with anti-CD28 and IL2 as above on days 3 and 6. On Day 9 PBMC were re-stimulated with peptide pools (2 µg/ml per peptide) and brefeldin A (10 µg/ml; Sigma-Aldrich). After 16-18 hours re-stimulation PBMC were harvested, washed in PBS, and stained for fixable live/dead (Near infrared, Thermo Fisher Scientific), washed in PBS, before being fixed in Fix/perm buffer (TF staining buffer kit, eBioscience) for 20 mins RT. Cells were washed in PBS and incubated in perm buffer (TF staining buffer kit, diluted 1:10 in ddH_2_O) for 20 mins RT, washed in PBS and resuspended in perm buffer with saturating concentrations of anti-human antibodies for intracellular staining: IL-2 PerCp-eFluor710 (Invitrogen, clone MQ1-17H12), TNFα FITC (BD bioscience, clone MAb11), CD8α BV785 (Biolegend, clone RPA-T8), IFNγ BV605 (BD biosciences, clone B27), IFNγ APC (Biolegend, clone 4S.B3), CD3 BUV805 (BD biosciences, clone UCHT1), CD4 BUV395 (BD biosciences, clone SK3), CD154 (CD40L) Pe-Cy7 (Biolegend, clone 24-31), MIP-1-β PE (BD biosciences, clone D21-1351). Cells were washed twice in PBS and analysed on a BD LSRII flow cytometer. Cytometer voltages were consistent across batches. FMOs and unstimulated samples were used to determine gates applied across samples. Data were analysed by FlowJo version 10.7 (TreeStar).

Optimisation experiments showed use of rhIL2 increases non-peptide specific proliferation of T cells but is essential for optimal expansion of proliferating cytokine producing peptide-specific T cells. CTV dilution and staining with anti-human-IFN**γ** antibodies was used to identify antigen-specific T cells. An unstimulated control well (equivalent DMSO to peptide wells added) was included for each PBMC sample and the percentage of CTV^lo^IFNγ+ CD4+ or CD8+ proliferating was subtracted from all peptide stimulated wells. T cell responses <0.1% of CD4 or CD8 T cells after 10-day peptide expansion or of less than 10 cells were excluded from analysis.

The T cell proliferation assay above was used to expand SARS-CoV-2-specific T cells and a 2-dimension matrix (**Supplementary Table 2**) was employed so that each 15mer peptide was represented in two pools aiding the identification of individuals immunogenic 15mer peptides. T cell responses were then confirmed by repeated expansion with individual 15mers.

### Sequence homology analyses

The sequence homology of SARS-CoV-2-derived peptides to HCoV sequences (**Fig. 3a**) was computed as previously described^40^. Briefly, the SARS-CoV-2 proteome (NC_045512.2) was decomposed into 15mer peptide sequences overlapping by 14 amino acids. A protein BLAST search of each 15mer peptide was then performed against a custom sequence database comprising 2531 *Coronaviridae* sequences^40^. Homology values of each SARS-CoV-2-derived peptide to viral accessions with ‘229E’, ‘OC43’, ‘NL63’, or ‘HKU1’ included in the species name and that were isolated from human hosts were retained (**Supplementary Table 1**). Additionally, to determine if the conservation of 15mer peptides differed between the SARS-CoV-2 proteins, the average homology of peptides within each protein was computed. A permutation test was conducted to test if the difference in average homology between the two proteins, *Δh*, was statistically significant. Briefly, the protein membership of each 15mer peptide was permuted (1000 iterations). The *Δh* of two proteins were then calculated at each iteration, resulting in a final null distribution of *Δh* values. *P*-values were computed as the number of permutations that yielded a *Δh* at least as extreme as the observed *Δh* of the two proteins. Custom scripts used to perform the homology searches, heatmap visualisation and permutation testing are hosted on GitHub (https://github.com/cednotsed/tcell_cross_reactivity_covid.git).

For sequence alignments of immunogenic 15mers or at described CD8 epitopes reference protein sequences for ORF1ab (accession numbers: QHD43415.1, NP_828849.2, YP_009047202.1, YP_009555238.1, YP_173236.1, YP_003766.2 and NP_073549.1) were downloaded from the NCBI database (https://www.ncbi.nlm.nih.gov/protein/) as previously described^10^. Sequences were aligned using the MUSCLE algorithm with default parameters and percentage identity was calculated in Geneious Prime 2020.1.2 (https://www.geneious.com). Alignment figures were made in Snapgene 5.1 (GSL Biotech).

### qPCR

Total RNA from Tempus blood was extracted using the Tempus Spin RNA isolation kit (Applied Biosystems, 4380204). cDNA was obtained using the High-Capacity cDNA Reverse Transcription Kit (Applied Biosystems). Quantitative PCR was performed using the TaqMan™ Fast Advanced Master Mix (Applied Biosystems) on ABI StepOnePlus Real-Time PCR machine (Applied Biosystems). The following cycling conditions were used: 95 °C for 2 mins, followed by 40 cycles of 95 °C for 3 s and 60 °C for 30 s. *IFI27* and *GAPDH* were amplified using the TaqMan Gene Expression Assay probes-Hs01086373_g1 (IFI27) and Hs02786624_g1 (GAPDH) respectively. *GAPDH* was used as a housekeeping gene control.

### Statistics and reproducibility

Data was assumed to have a non-Gaussian distribution and nonparametric tests were used throughout. For single paired and unpaired comparisons Wilcoxon matched-pairs signed rank test and a Mann-Whitney U test were used. For multiple unpaired comparisons Kruskal-Wallis one-way ANOVA with Dunn’s correction was used. For correlations, Spearman’s r test was used. A p value <0.05 was considered significant. Prism v. 7.0e and 8.0 for Mac was used for analysis. Details are provided in figure legends.

### Data reporting

Power calculations were used to estimate the sample size needed for week 16 sub study (see above). No statistical methods were used to predetermine sample size. For all assays samples from each cohort were run in parallel to reduce the impact of inter-batch technical variation.

IFNγ-ELISpot assays were performed on HCW cohorts prior to unblinding of group (Laboratory-confirmed-infection or exposed seronegative). Other experiments were not randomized and the investigators were not blinded to allocation during experiments and outcome assessment.

## Data availability statements

All data analysed during this study are included in this published article (and its supplementary information files). Custom scripts used to perform the homology searches, heatmap visualisation and permutation testing are hosted on GitHub (https://github.com/cednotsed/tcell_cross_reactivity_covid.git). The datasets generated during and/or analysed during the current study are available from the corresponding author on reasonable request. Correspondence and requests for materials should be addressed to MKM or LS.

## Acknowledgments

We are extremely grateful to all patients and control volunteers who participated in this study and to all clinical staff who helped with recruitment and sample collection. We are grateful to Jamie Evans at the Rayne Building FACS facility for assistance with Flow cytometry assays.

## Author contributions

MKM conceived the project and obtained funding. LS, MN, AB and MKM designed experiments. CM, TAT, JCM, MN, ÁM established the HCW cohort. LS, MOD, NMS, OEA, CP, JMG, SK, GAM, JR, JD, GJ, collected or processed HCW samples with COVIDsortium investigators. MKM, LS, MN, and ESW established medical student/laboratory staff and pre-pandemic cohorts (UK). LM, MJ, DG and COVIDsortium investigators performed serology. MKM and LS designed T cell experiments. LS, MOD, NMS, OEA, developed, performed and analysed the T cell experiments. AB, NLB, ATT, CYLT performed T cell assays and analysed data from pre-pandemic cohort (Singapore). AC performed and analysed blood transcriptomic experiments. AMV supervised HLA analysis. ÁM supervised nAb experiments. JMG and CP performed and analysed nAb experiments. CT, FB performed viral sequence analysis. ESW, MPJ, GJ, RJB, CM, TAT, JCM, AM, MD, provided or processed essential clinical data. LS, AC, CP, JMG, NLB, AT, CT, AMV, DMA, RJB, CM, TAT, LM, FB, AM, MN, AB, and MKM analysed and interpreted the data. LS and MKM prepared the manuscript. All authors provided critical review of the manuscript.

## UK COVIDsortium Investigators

Hakam Abbass^4^, Mashael Alfarih^4^, Zoe Alldis^4^, Daniel M Altmann^10^, Oliver E Amin^7^, Mervyn Andiapen^4^, Jessica Artico^4^, João B Augusto^4^, Georgina L Baca^4^, Sasha N L. Bailey^1^, Anish N Bhuva^4^, Alex Boulter^4^, Ruth Bowles^4^, Rosemary J Boyton^1^, Olivia V Bracken^12^, Ben O’Brien^4^, Tim Brooks^3^, Natalie Bullock^2^, David K Butler^1^, Gabriella Captur^5,8^, Nicola Champion^4^, Carmen Chan^4^, Aneesh Chandran^7^, Jorge Couto de Sousa^4^, Xose Couto-Parada^4^, Teresa Cutino-Moguel^4^, Rhodri H Davies^4^, Brooke Douglas^5^, Cecilia Di Genova^13^, Keenan Dieobi-Anene^4^, Mariana O Diniz^7^, Anaya Ellis^3^, Karen Feehan^12^, Malcolm Finlay^4^, Marianna Fontana^5^, Nasim Forooghi^4^, Joseph M Gibbons^2^, Derek Gilroy^14^, Matt Hamblin^4^, Gabrielle Harker^3^, Jacqueline Hewson^3^, Lauren M Hickling^15^, Aroon D Hingorani^7^, Lee Howes^8^, Ivie Itua^4^, Victor Jardim^4^, Wing-Yiu Jason Lee^2^, Melanie petra Jensen^4^, Jessica Jones^3^, Meleri Jones^2^, George Joy^4^, Vikas Kapil^4,16^, Hibba Kurdi^4,8^, Jonathan Lambourne^4^, Kai-Min Lin^1^, Sarah Louth^5^, Mala K Maini^7^, Vineela Mandadapu^4^, Charlotte Manisty^4,8^, Áine McKnight^2^, Katia Menacho^4^, Celina Mfuko^4^, Oliver Mitchelmore^4^, Christopher Moon^3^, James C Moon^4,8^, Diana Munoz-Sandoval^1’^ Sam M Murray^1^, Mahdad Noursadeghi^7^, Ashley Otter^3^, Corinna Pade^2^, Susana Palma^4^, Ruth Parker^17^, Kush Patel^4^, Babita Pawarova^5^, Steffen E Petersen^4^, Brian Piniera^4^, Franziska P Pieper^1^, Lisa Rannigan^5^, Alicja Rapala^8^, Catherine J Reynolds^1^, Amy Richards^4^, Matthew Robathan^16^, Joshua Rosenheim^7^, Genine Sambile^4^, Nathalie M. Schmidt^7^, Amanda Semper^3^, Andreas Seraphim^4^, Mihaela Simion^5^, Angelique Smit^5^, Michelle Sugimoto^12^, Leo Swadling^7^, Stephen Taylor^3^, Nigel Temperton^13^, Stephen Thomas^3^, George D Thornton^4,8^, Thomas A Treibel^4,8^, Art Tucker^4^, Jessry Veerapen^4^, Mohit Vijayakumar^4^, Sophie Welch^4^, Theresa Wodehouse^4^, Lucinda Wynne^4^, Dan Zahedi^17^

^1^Department of Infectious Disease, Imperial College London, London, UK.

^2^Blizard Institute, Barts and the London School of Medicine and Dentistry, Queen Mary University of London, London, UK.

^3^National Infection Service, Public Health England, Porton Down, UK.

^4^St Bartholomew’s Hospital, Barts Health NHS Trust, London, UK.

^5^Royal Free London NHS Foundation Trust, London, UK.

^6^James Wigg Practice, Kentish Town, London, UK.

^7^Division of Infection and Immunity, University College London, London, UK.

^8^Institute of Cardiovascular Science, University College London, UK.

^9^Academic Rheumatology, Clinical Sciences, Nottingham City Hospital, Nottingham, UK.

^10^Department of Immunology and Inflammation, Imperial College London, London, UK.

^11^Lung Division, Royal Brompton and Harefield Hospitals, London, UK.

^12^ Division of Medicine, University College London, London, UK.

^13^Viral Pseudotype Unit, Medway School of Pharmacy, Chatham Maritime, Kent, UK.

^14^ Centre for Clinical Pharmacology, University College London, London, UK.

^15^East London NHS Foundation Trust Unit for Social and Community Psychiatry, Newham Centre for Mental Health, London, UK.

^16^William Harvey Research Institute, Queen Mary University of London, London, UK.

^17^ School of Clinical Medicine, University of Cambridge, Cambridge, UK.

## Extended Data

**Extended Data Fig. 1.**
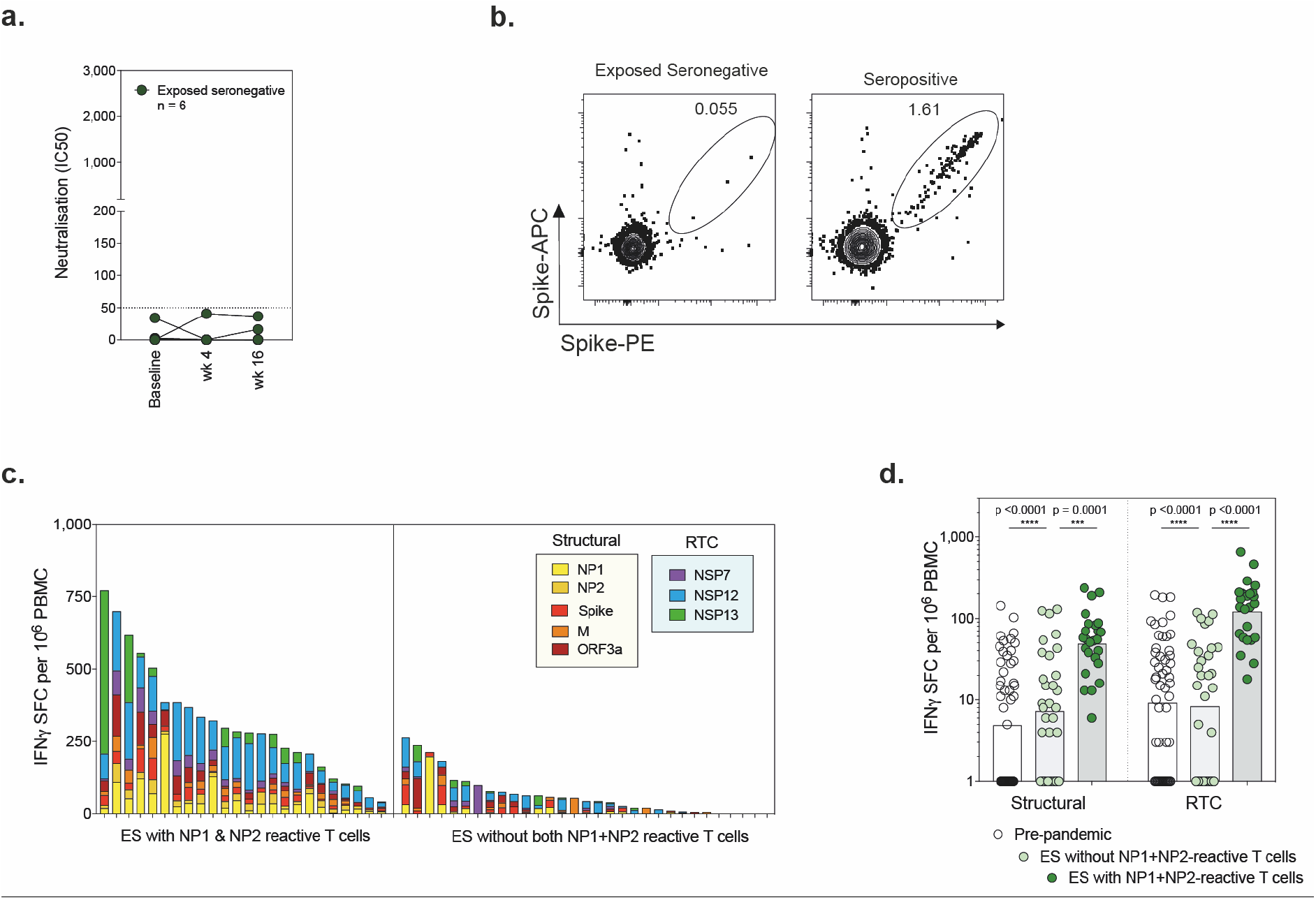
SARS-CoV-2 immunity in exposed seronegative healthcare workers – authentic virus (Wuhan Hu-1) neutralisation and T cell response in those with NP1+NP2 responses. **a**, authentic virus neutralisation at 3 time-points, n=6. **b**, Example plots of SARS-COV-2 spike memory B cell staining (gated on: lymphocytes, singlets, Live, CD3-CD14-CD19+, CD20+, excluding CD38hi, IgD+ and CD21+CD27-fractions) in an exposed seronegative HCW at wk16 and a seropositive individual. **c**, Magnitude of T cell response coloured by viral protein in ES with T cells reactive against both NP1 and NP (left) and against one of or neither NP1 or NP2 pools (right) at wk16. **d**, Summed response to RTC and structural regions of SARS-CoV-2 in pre-pandemic samples and ES with and without NP1+NP2-reactive T cell responses at wk16. Kruskal-Wallis with Dunn’s correction. Bars, geomean. NP, nucleoprotein.

**Extended Data Fig. 2.**
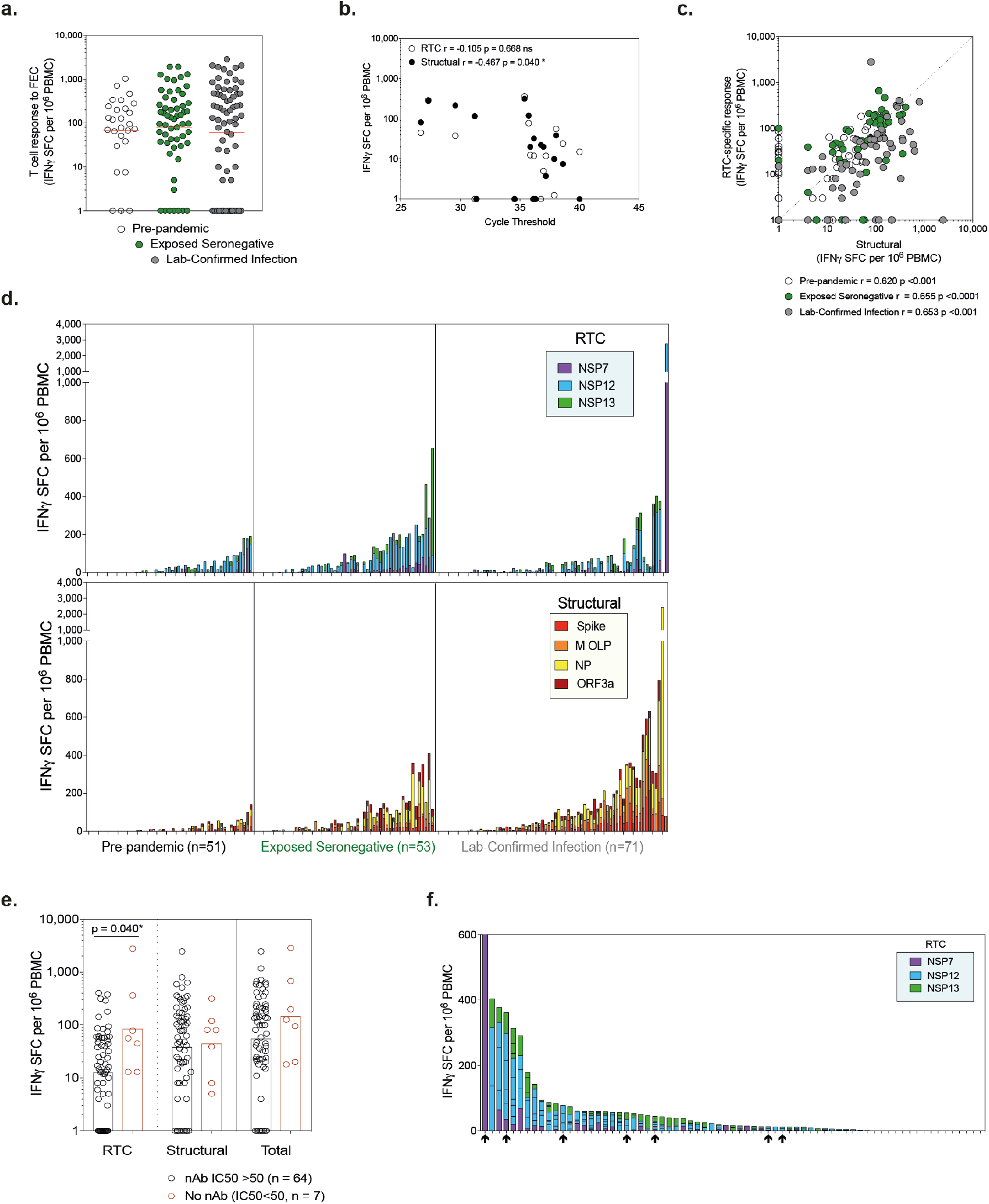
T cell responses to RTC and Structural regions of SARS-CoV-2 by cohort: **a**, T cell response to Flu, EBV and CMV (FEC) MHC class I restricted peptide pool. **b**, E gene RT-PCR cycle threshold value vs. magnitude of T cell response to RTC or Structural proteins in HCW with laboratory-confirmed infection. **c**, Magnitude of T cell response to RTC vs. structural regions. **d**, Magnitude of T cell response to RTC (top) and structural regions (bottom) coloured by specificity. **e**, Magnitude of T cell response in laboratory-confirmed infected group in HCW with or without detectable nAb at wk16. **f**, T cell response to RTC coloured by protein in laboratory-confirmed infected group ordered by magnitude. HCW lacking neutralising antibodies highlighted by arrows below. **a-f**, IFN**γ**-ELISpot wk16. **a**,**e**, Bars at geomean. **b-c** Spearman r. **a**,**e** Kruskal-Wallis ANOVA with Dunn’s correction.

**Extended Data Fig. 3.**
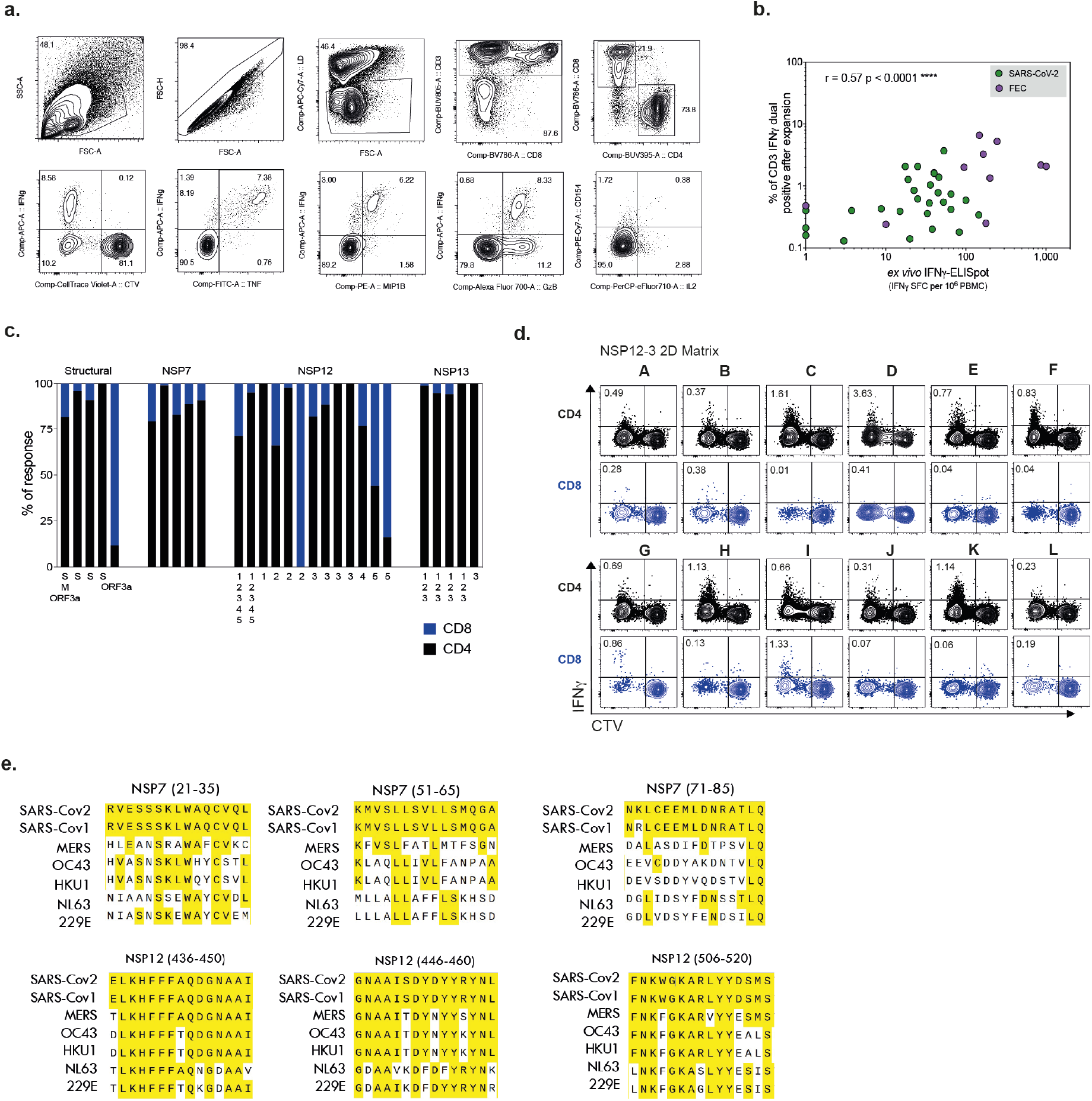
Functional and proliferative SARS-CoV-2 specific T cells in exposed seronegative HCW. **a**, example gating of CTV stained PBMC after 10-day peptide stimulation: Lymphocytes (SSC-A vs. FSC-A), single cells (FSC-H vs. FSC-A), Live cells (fixable live/dead-), CD3+, CD4+ or CD8+. Second row: Gated on CD8+ showing cytokine combinations. Response to immunodominant MHC class I-restricted peptide pool against Flu, EBV, CMV (FEC) in exposed seronegative HCW. **b**, Correlation between the magnitude of T cells responses to SARS-CoV-2 pools or FEC after 10-day *in vitro* expansion (% dual staining for two anti-human IFN mAb clones, unexpanded responses <0.1% of CD3 post-expansion excluded) and *ex vivo* IFN -ELISpot in exposed seronegative HCW. Spearman r. **c**, Proportion of SARS-CoV-2-specific T cells (CTV-IFN +) that are CD4+ or CD8+ after 10-day expansion (where sub pools were used, they are indicated below bar). **d**, Example 2D-mapping matrix after 10-day expansion with NSP12-3 peptide pool in an exposed seronegative HCW (Antigen-specific identified as CTV^lo^IFN -APC+). **e**, Alignment of *Coronaviridae* consensus sequences at immunogenic 15mers peptides. Conserved amino acids in yellow. **a-d** ES at wk16.

**Extended Data Fig. 4.**
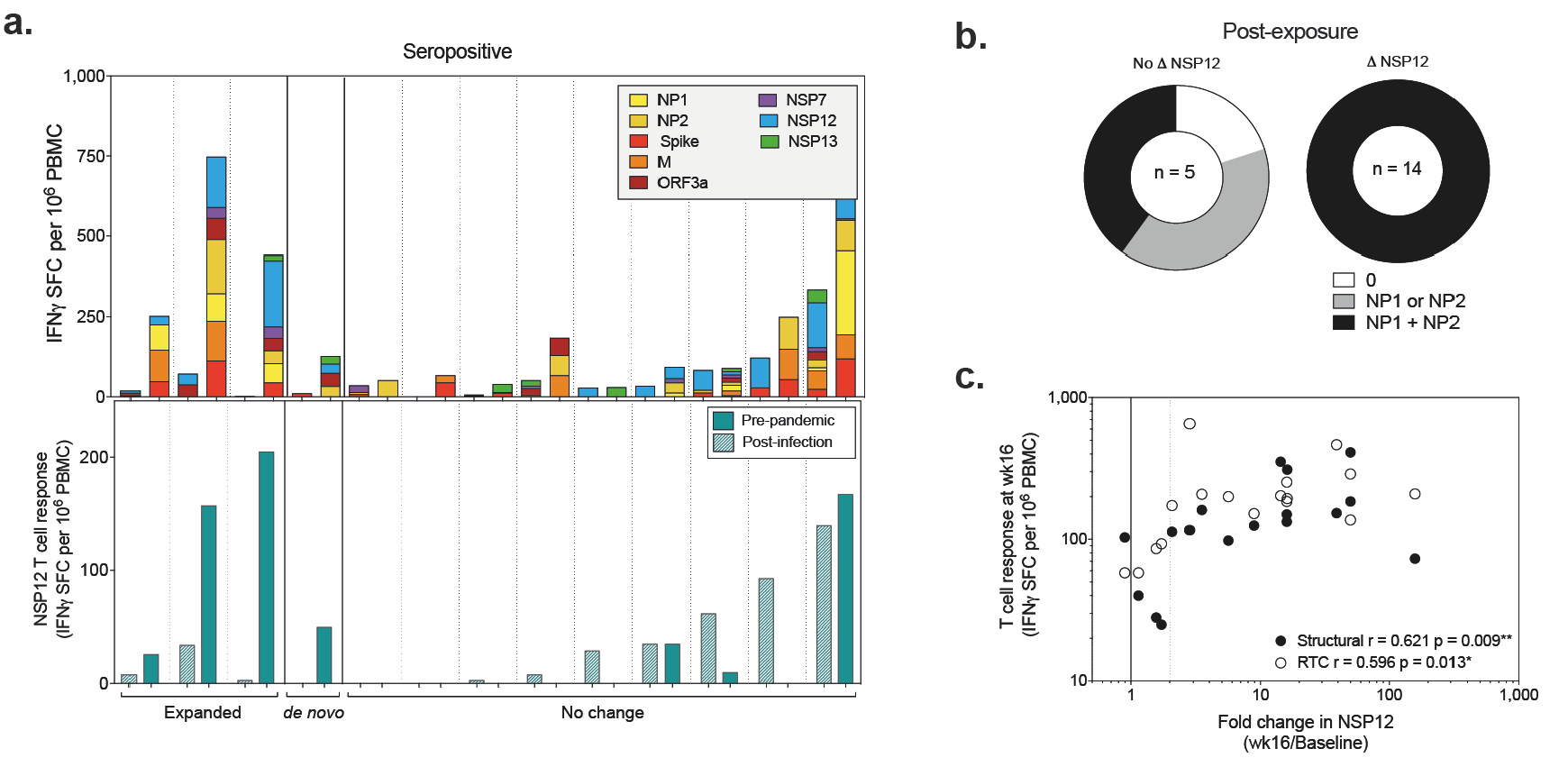
*In vivo* expansion of pre-existing SARS-CoV-2-reactive T cells post-infection or post-exposure. **a**, Change in magnitude of T cell response between pre-pandemic and post-infection samples (upper panel: all proteins, lower panel: NSP12) from seropositive medical students and laboratory staff. **b**, Proportion of ES with NP1 + NP2-reactive T cells grouped by those with and without *de novo* or expanded NSP12 responses at wk16. **c**, Correlation between the fold-change in NSP12 between recruitment and wk16 and total response to RTC or structural proteins at wk16 in ES. Spearman r.

**Extended Data Table 1.** Cohort Demographics

|  | Lab Confirmed infection | Seronegatives | Pre-pandemic (London) | Pre-pandemic (Singapore) |
| --- | --- | --- | --- | --- |
| **Number of subjects** | 76 | 57 | 53 | 12* |
| **Mean age (range)** | 41.7 (25-62) | 37.7 (21-62) | 26.5 (20-59) | 40 (30-59) |
| **Gender:** |  |  |  |  |
| Female, n (%) | 50 (65.79) | 35 (61.40) | 35 (66.04) | 4 (33.33) |
| Male, n (%) | 26 (34.21) | 22 (38.60) | 18 (33.96) | 8 (66.67) |
| **Ethnicity:** |  |  |  |  |
| White, n (%) | 55 (72.37) | 40 (70.18) | 39 (73.58) | 2 (16.67) |
| Other, n (%) | 21 (27.63) | 17 (29.82) | 14 (26.42) | 10 (83.33) |
| * demographics for 4 pre-pandemic samples unknown |  |  |  |  |

**Extended Data Table 2:** CTV T cell proliferation in exposed seronegative HCW.

| Donor | Week | Antigen | Subpool | Minipool | Peptide | CD4 % CTV-IFNγ | CD8 % CTV-IFNγ |
| --- | --- | --- | --- | --- | --- | --- | --- |
| 1 | 16 | FEC | - | - | - | 0.14% | 5.33% |
| 2 | 16 | NSP7 | - | A&G | 9 | 0.27% | 0.42% |
| 2 | 16 | NSP12 | 3 | E&G | 79 | 0.61% | 0.54% |
| 2 | 16 | NSP13 | 3 | E&M | 118 | 0.23% | 0.14% |
| 2 | 16 | NSP12 | 5 | E&J | 171 | 0.13% | 0.26% |
| 2 | 16 | NSP7 | - | A&F | 5 | 0.39% | 0.53% |
| 3 | 16 | Spike/M/ORF3a | - | - | - | 0.11% | 0.02% |
| 3 | 16 | FEC | - | - | - | 0.00% | 10.99% |
| 3 | 16 | NSP7 | - | - | - | 0.81% | 0.01% |
| 3 | 16 | NSP12 | - | - | - | 0.23% | 0.01% |
| 3 | 16 | NSP13 | - | - | - | 1.04% | 0.06% |
| 3 | 16 | NSP7 | - | A | - | 0.36% | 0.01% |
| 3 | 16 | NSP7 | - | B | - | 0.49% | 0.01% |
| 3 | 16 | NSP7 | - | C | - | 1.23% | 0.00% |
| 3 | 16 | NSP7 | - | D | - | 0.53% | 0.00% |
| 3 | 16 | NSP7 | - | E | - | 0.68% | 0.00% |
| 3 | 16 | NSP7 | - | F | - | 0.49% | 0.01% |
| 3 | 16 | NSP7 | - | G | - | 1.12% | 0.02% |
| 3 | 16 | NSP7 | - | H | - | 1.57% | 0.01% |
| 3 | 16 | NSP7 | - | C&G | 11 | 0.30% | 0.00% |
| 3 | 16 | NSP7 | - | C&H | 15 | 0.11% | 0.01% |
| 3 | 16 | NSP7 | - | A&F | 5 | 0.16% | 0.00% |
| 3 | 16 | NSP12 | 3 | B&I | 88 | 0.07% | 0.00% |
| 3 | 16 | NSP7 | - | A&F | 5 | 0.01% | 0.06% |
| 3 | 16 | NSP7 | - | - | - | 0.18% | 0.12% |
| 3 | 16 | NSP12 | 3 | I | - | 0.01% | 0.01% |
| 3 | 16 | NSP12 | 3 | - | - | 0.16% | 0.02% |
| 3 | 16 | NSP12 | 3 | B&G | 76 | 0.00% | 0.01% |
| 3 | 16 | NSP12 | 3 | B&I | 88 | 0.02% | 0.00% |
| 3 | 16 | NSP12 | 3 | D&I | 90 | 0.01% | 0.00% |
| 4 | 16 | NSP12 | 4 | - | 148 | 0.10% | 0.02% |
| 4 | 16 | NSP12 | 5 | E&J | 171 | 0.17% | 0.01% |
| 4 | 16 | NSP12 | 3 | - | - | 0.09% | 0.00% |
| 5 | 16 | FEC | - | - | - | 1.14% | 0.23% |
| 5 | 16 | NSP7 | - | - | - | 0.00% | 0.00% |
| 5 | 16 | NSP12 | - | - | - | 2.07% | 0.45% |
| 5 | 16 | NSP13 | - | - | - | 0.58% | 0.04% |
| 5 | 16 | Spike | - | - | - | 0.20% | 0.01% |
| 5 | 16 | NSP12 | 3 | A | - | 1.68% | 0.31% |
| 5 | 16 | NSP12 | 3 | B | - | 0.38% | 0.13% |
| 5 | 16 | NSP12 | 3 | C | - | 0.84% | 0.23% |
| 5 | 16 | NSP12 | 3 | D | - | 1.12% | 0.26% |
| 5 | 16 | NSP12 | 3 | E | - | 0.77% | 0.34% |
| 5 | 16 | NSP12 | 3 | F | - | 1.36% | 0.40% |
| 5 | 16 | NSP12 | 3 | G | - | 1.26% | 0.20% |
| 5 | 16 | NSP12 | 3 | H | - | 0.67% | 0.07% |
| 5 | 16 | NSP12 | 3 | I | - | 1.81% | 0.23% |
| 5 | 16 | NSP12 | 3 | J | - | 0.37% | 0.29% |
| 5 | 16 | NSP12 | 3 | K | - | 0.60% | 0.29% |
| 5 | 16 | NSP12 | 3 | L | - | 0.65% | 0.44% |
| 5 | 16 | NSP12 | 3 | A&I | 87 | 0.00% | 0.00% |
| 5 | 16 | NSP12 | 3 | F&I | 92 | 0.00% | 0.00% |
| 6 | 16 | FEC | - | - | - | 0.07% | 8.56% |
| 6 | 16 | NP | 1&2 | - | - | 0.00% | 0.19% |
| 6 | 16 | ORF3a | - | - | - | 0.00% | 0.00% |
| 6 | 16 | Spike/M/ORF3a | - | - | - | 0.00% | 0.00% |
| 6 | 16 | NSP7 | - | - | - | 0.61% | 0.08% |
| 6 | 16 | NSP12 | - | - | - | 0.00% | 0.13% |
| 6 | 16 | NSP13 | - | - | - | 0.30% | 0.00% |
| 6 | 16 | NSP12 | 1 | - | - | 0.81% | 0.00% |
| 6 | 16 | NSP12 | 2 | - | - | 0.11% | 0.06% |
| 6 | 16 | NSP12 | 3 | - | - | 0.62% | 0.08% |
| 6 | 16 | NSP12 | 3 | - | - | 0.03% | 0.14% |
| 6 | 16 | NSP12 | 3 | A | - | 0.36% | 0.26% |
| 6 | 16 | NSP12 | 3 | B | - | 0.24% | 0.36% |
| 6 | 16 | NSP12 | 3 | C | - | 1.48% | 0.00% |
| 6 | 16 | NSP12 | 3 | D | - | 3.50% | 0.39% |
| 6 | 16 | NSP12 | 3 | E | - | 0.64% | 0.02% |
| 6 | 16 | NSP12 | 3 | F | - | 0.70% | 0.02% |
| 6 | 16 | NSP12 | 3 | G | - | 0.56% | 0.84% |
| 6 | 16 | NSP12 | 3 | H | - | 1.00% | 0.11% |
| 6 | 16 | NSP12 | 3 | I | - | 0.53% | 1.31% |
| 6 | 16 | NSP12 | 3 | J | - | 0.18% | 0.05% |
| 6 | 16 | NSP12 | 3 | K | - | 1.01% | 0.04% |
| 6 | 16 | NSP12 | 3 | L | - | 0.10% | 0.17% |
| 6 | 16 | NSP12 | 3 | B&G | 76 | 0.00% | 0.00% |
| 6 | 16 | NSP12 | 3 | D&H | 84 | 0.05% | 0.00% |
| 6 | 16 | NSP12 | 3 | B&I | 88 | 0.23% | 0.11% |
| 6 | 16 | NSP12 | 3 | D&K | 102 | 0.08% | 0.00% |
| 6 | 16 | NSP12 | 5 | E&J | 171 | 0.11% | 0.03% |
| 7 | 16 | FEC | - | - | - | 5.00% | 12.73% |
| 8 | 16 | FEC | - | - | - | 0.76% | 4.48% |
| 8 | 16 | NP | 1&2 | - | - | 0.01% | 0.01% |
| 8 | 16 | NSP12 | 1/2/3 | - | - | 0.00% | 0.00% |
| 8 | 16 | ORF3a | - | - | - | 0.07% | 0.52% |
| 8 | 16 | Spike/M/ORF3a | - | - | - | 0.04% | 0.14% |
| 9 | 16 | NSP7 | - | A&F | 5 | 0 | 0.06% |
| 10 | na | NSP12 | - | - | - | 1.47% | 0.60% |
| 10 | na | NSP12 | 5 | - | - | 0.21% | 0.27% |
| 10 | na | NSP13 | - | - | - | 3.40% | 0.04% |
| 10 | na | Spike | - | - | - | 0.10% | 0.00% |
| 10 | na | NSP12 | 3 | A | - | 0.16% | 0.41% |
| 10 | na | NSP12 | 3 | B | - | 0.18% | 0.10% |
| 10 | na | NSP12 | 3 | C | - | 0.08% | 0.12% |
| 10 | na | NSP12 | 3 | D | - | 0.27% | 0.25% |
| 10 | na | NSP12 | 3 | E | - | 0.28% | 0.30% |
| 10 | na | NSP12 | 3 | F | - | 0.09% | 0.28% |
| 10 | na | NSP12 | 3 | G | - | 0.32% | 0.00% |
| 10 | na | NSP12 | 3 | H | - | 0.17% | 0.15% |
| 10 | na | NSP12 | 3 | I | - | 0.21% | 1.71% |
| 10 | na | NSP12 | 3 | J | - | 0.16% | 0.06% |
| 10 | na | NSP12 | 3 | K | - | 0.27% | 0.18% |
| 10 | na | NSP12 | 3 | L | - | 0.10% | 0.37% |
| 10 | na | NSP12 | 3 | - | - | 1.61% | 0.00% |
| 10 | na | NSP12 | 3 | I | - | 0.13% | 0.03% |
| 10 | na | NSP12 | 3 | A&I | 87 | 0.00% | 0.00% |
| 10 | na | NSP12 | 3 | B&I | 88 | 0.10% | 0.02% |
| 10 | na | NSP12 | 3 | C&I | 89 | 0.00% | 0.00% |
| 10 | na | NSP12 | 3 | D&I | 90 | 0.00% | 0.08% |
| 10 | na | NSP12 | 3 | D&I | 90 | 0.17% | 0.03% |
| 10 | na | NSP12 | 3 | E&I | 91 | 0.00% | 0.00% |
| 10 | na | NSP12 | 3 | F&I | 92 | 0.00% | 0.00% |
| 10 | na | NSP7 | - | A&F | 5 | 0.78% | 0.00% |
| 10 | na | NSP12 | 3 | B&G | 76 | 0.00% | 0.00% |
| 10 | na | NSP12 | 3 | E&G | 79 | 0.00% | 0.00% |

**Extended Data Table 3.**
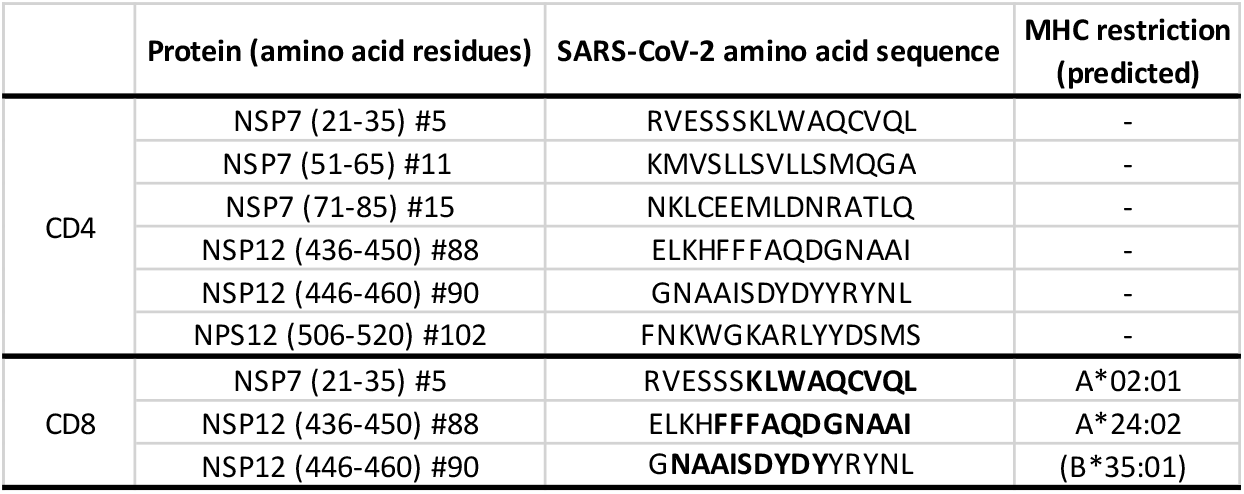
Immunogenic peptides recognised by CD4+ or CD8+ T cells in ES at wk16.

**Extended Data Table 4.** Demographics and sampling of medical student/Laboratory staff cohort.

|  | Serostatus/PCR | Gender | Age (post exposure sample) | Exposure | NSP12 Response | Post-exposure sample: Months since known exposure/PCR+ or symptoms |
| --- | --- | --- | --- | --- | --- | --- |
| Close contact | Seronegative | F | 20-24 | Household Contact | - | 8-9 |
|  | Seronegative | M | 20-24 | Suspected household contact* | Expanded | 8-9 |
|  | Seronegative | M | 20-24 | Close Contact | de novo | 7-8 |
|  | Seronegative | F | 20-24 | Suspected household contact* | de novo | 6-7 |
|  | Seronegative | M | 20-24 | Suspected household contact* | de novo | 9-10 |
|  | Seronegative | F | 20-24 | Close Contact | Expanded | 9-10 |
|  | Seronegative | F | 20-24 | Household Contact | de novo | 9-10 |
|  | Seronegative | M | 20-24 | Close Contact | Expanded | 8-9 |
|  | Seronegative | M | 65-69 | Household Contact | Expanded | 7-8 |
|  | Seronegative | F | 25-29 | Household Contact | - | 6-7 |
| Lab confirmed infection | Seropositive | M | 20-24 | not known | - | 9-10 |
|  | Seropositive | F | 20-24 | Household Contact | - | 2-3 |
|  | Seropositive | F | 20-24 | Close Contact | - | 9-10 |
|  | Seropositive | F | 20-24 | Suspected household contact* | - | 9-10 |
|  | PCR+ Seropositive | F | 20-24 | Close Contact | de novo | 2-3 |
|  | PCR+ Seropositive | M | 20-24 | Close Contact | Expanded | 1-2 |
|  | Seropositive | F | 20-24 | Close Contact | Expanded | 9-10 |
|  | PCR+ Seropositive | F | 20-24 | Household Contact | - | 1-2 |
|  | Seropositive | F | 25-29 | not known | Expanded | 4-5 |
|  | Seropositive | M | 25-29 | Household Contact | - | 5-6 |
|  | Seropositive | F | 25-29 | not known | - | 4-5 |
|  | PCR+ Seropositive | F | 30-34 | Occupational | - | 1-2 |
|  | PCR+ Seropositive | M | 25-29 | not known | - | 1-2 |

\* Household contact with case-defining symptoms but no PCR confirmation
Expanded = $\geq 2$ fold or $>35$ SFU/ $10^6$ PBMC increase in NSP12 response from pre-pandemic to post-exposure or infection time point

## **Supplementary Tables** (Separate excel file submitted)

**Supplementary Table 1:** HCoV sequence accession numbers

**Supplementary Table 2:** 2D epitope mapping matrices.

**Supplementary Table 3:** NSP12 Overlapping peptide sequences.

## References

1. Tan, A. T. et al. Early induction of functional SARS-CoV-2-specific T cells associates with rapid viral clearance and mild disease in COVID-19 patients. Cell Rep. 34, 108728 (2021).

2. Zhao, J. J. et al. Airway Memory CD4+ T Cells Mediate Protective Immunity against Emerging Respiratory Coronaviruses. Immunity 44, 1379–1391 (2016).

3. McMahan, K. et al. Correlates of protection against SARS-CoV-2 in rhesus macaques. Nature 590, 630–634 (2021).

4. Sun, J. et al. Generation of a Broadly Useful Model for COVID-19 Pathogenesis, Vaccination, and Treatment. Cell 182, 734-743.e5 (2020).

5. Rydyznski Moderbacher, C. et al. Antigen-Specific Adaptive Immunity to SARS-CoV-2 in Acute COVID-19 and Associations with Age and Disease Severity. Cell 183, 996-1012.e19 (2020).

6. Sekine, T. et al. Robust T Cell Immunity in Convalescent Individuals with Asymptomatic or Mild COVID-19. Cell 183, 158-168.e14 (2020).

7. Mateus, J. et al. Selective and cross-reactive SARS-CoV-2 T cell epitopes in unexposed humans. Science (80-.). 370, 89–94 (2020).

8. Nelde, A. et al. SARS-CoV-2-derived peptides define heterologous and COVID-19-induced T cell recognition. Nat. Immunol. 22, 74–85 (2021).

9. Braun, J. et al. SARS-CoV-2-reactive T cells in healthy donors and patients with COVID-19. Nature 587, 270–274 (2020).

10. Le Bert, N. et al. SARS-CoV-2-specific T cell immunity in cases of COVID-19 and SARS, and uninfected controls. Nature 584, 457–462 (2020).

11. Grifoni, A. et al. Targets of T Cell Responses to SARS-CoV-2 Coronavirus in Humans with COVID-19 Disease and Unexposed Individuals. Cell 181, 1489-1501.e15 (2020).

12. Schulien, I. et al. Characterization of pre-existing and induced SARS-CoV-2-specific CD8+ T cells. Nat. Med. 27, 78–85 (2021).

13. Perlman, S. & Netland, J. Coronaviruses post-SARS: Update on replication and pathogenesis. Nat. Rev. Microbiol. 7, 439–450 (2009).

14. De Wit, E., Van Doremalen, N., Falzarano, D. & Munster, V. J. SARS and MERS: Recent insights into emerging coronaviruses. Nat. Rev. Microbiol. 14, 523–534 (2016).

15. Yamada, T. et al. RIG-I triggers a signaling-abortive anti-SARS-CoV-2 defense in human lung cells. Nat. Immunol. (2021) doi:10.1038/s41590-021-00942-0.

16. Gupta, R. K. et al. Blood transcriptional biomarkers of acute viral infection for detection of pre-symptomatic SARS-CoV-2 infection. medRxiv 2021.01.18.21250044 (2021) doi:10.1101/2021.01.18.21250044.

17. Wang, Z. et al. Exposure to SARS-CoV-2 generates T-cell memory in the absence of a detectable viral infection. Nat. Commun. 12, 1724 (2021).

18. Ogbe, A. et al. T cell assays differentiate clinical and subclinical SARS-CoV-2 infections from cross-reactive antiviral responses. Nat. Commun. 12, (2021).

19. da Silva Antunes, R. et al. Differential T cell reactivity to endemic coronaviruses and SARS-CoV-2 in community and health care workers. J. Infect. Dis. 2021.01.12.21249683 (2021) doi:10.1093/infdis/jiab176.

20. Gallais, F. et al. Intrafamilial exposure to SARS-CoV-2 associated with cellular immune response without Seroconversion, France. Emerg. Infect. Dis. 27, 113–121 (2021).

21. Reynolds, C. J. et al. Discordant neutralizing antibody and T cell responses in asymptomatic and mild SARS-CoV-2 infection. Sci. Immunol. 5, (2020).

22. Low, J. S. et al. Clonal analysis of immunodominance and cross-reactivity of the CD4 T cell response to SARS-CoV-2. Science (80-.). eabg8985 (2021) doi:10.1126/science.abg8985.

23. Bonifacius, A. et al. COVID-19 immune signatures reveal stable antiviral T cell function despite declining humoral responses. Immunity 54, 340-354.e6 (2021).

24. Saini, S. K. et al. SARS-CoV-2 genome-wide T cell epitope mapping reveals immunodominance and substantial CD8 ^+^ T cell activation in COVID-19 patients. Sci. Immunol. 6, eabf7550 (2021).

25. Ferretti, A. P. et al. Unbiased Screens Show CD8+ T Cells of COVID-19 Patients Recognize Shared Epitopes in SARS-CoV-2 that Largely Reside outside the Spike Protein. Immunity 53, 1095-1107.e3 (2020).

26. Lizeng, Q. et al. Potent Neutralizing Serum Immunoglobulin A (IgA) in Human Immunodeficiency Virus Type 2-Exposed IgG-Seronegative Individuals. J. Virol. 78, 7016–7022 (2004).

27. Ritchie, A. J. et al. Differences in HIV-Specific T Cell Responses between HIV-Exposed and -Unexposed HIV-Seronegative Individuals. J. Virol. 85, 3507–3516 (2011).

28. Wiegand, J. et al. HBV-specific T-cell responses in healthy seronegative sexual partners of patients with chronic HBV infection. J. Viral Hepat. 17, 631–639 (2010).

29. Tsertsvadze, T. et al. The natural history of recent hepatitis C virus infection among blood donors and injection drug users in the country of Georgia Positive-strand RNA viruses. Virol. J. 13, 1–7 (2016).

30. Rivino, L. et al. Differential Targeting of Viral Components by CD4+ versus CD8+ T Lymphocytes in Dengue Virus Infection. J. Virol. 87, 2693–2706 (2013).

31. Heller, T. et al. Occupational exposure to hepatitis C virus: Early T-cell responses in the absence of seroconversion in a longitudinal cohort study. J. Infect. Dis. 208, 1020– 1025 (2013).

32. Rowland-Jones, S. et al. HIV-specific cytotoxic T-cells in HIV-exposed but uninfected Gambian women. Nat. Med. 1, 59–64 (1995).

33. Turtle, L. et al. Human T cell responses to Japanese encephalitis virus in health and disease. J. Exp. Med. 213, 1331–1352 (2016).

34. Post, J. J. et al. Clearance of hepatitis C viremia associated with cellular immunity in the absence of seroconversion in the hepatitis C incidence and transmission in prisons study cohort. J. Infect. Dis. 189, 1846–1855 (2004).

35. Promadej, N., Costello, C. & Wernett, M. M. Erratum: Broad human immunodeficiency virus (HIV)-specific T cell responses to conserved HIV proteins in HIV-seronegative women highly exposed to a single HIV-infected partner (Journal of Infectious Diseases (2003) 187 (1053-1063)). J. Infect. Dis. 187, 1346 (2003).

36. Manisty, C. et al. Time series analysis and mechanistic modelling of heterogeneity and sero-reversion in antibody responses to mild SARS CoV-2 infection. EBioMedicine 65, 103259 (2021).

37. Jeffery-Smith, A. et al. SARS-CoV-2-specific memory B cells can persist in the elderly despite loss of neutralising 1 antibodies 2 3 4. bioRxiv 2021.05.30.446322 (2021) doi:10.1101/2021.05.30.446322.

38. Chandran, A. et al. Title Non-severe SARS-CoV-2 infection is characterised by very early T cell proliferation independent of type 1 interferon responses and distinct from other acute respiratory viruses. medRxiv 2021.03.30.21254540 (2021) doi:10.1101/2021.03.30.21254540.

39. Kalimuddin, S. et al. Early T cell and binding antibody responses are associated with COVID-19 RNA vaccine efficacy onset. Med 0, 1–7 (2021).

40. Tan, C. C. S. et al. Pre-existing T cell-mediated cross-reactivity to SARS-CoV-2 cannot solely be explained by prior exposure to endemic human coronaviruses 2 3. bioRxiv 2020.12.08.415703 (2020).

41. Lineburg, K. E. et al. CD8+ T cells specific for an immunodominant SARS-CoV-2 nucleocapsid epitope cross-react with selective seasonal coronaviruses. Immunity (2021) doi:10.1016/j.immuni.2021.04.006.

42. Tarke, A. et al. Comprehensive analysis of T cell immunodominance and immunoprevalence of SARS-CoV-2 epitopes in COVID-19 cases. Cell Reports Med. 2, (2021).

43. Treibel, T. A. et al. COVID-19: PCR screening of asymptomatic health-care workers at London hospital. Lancet 395, 1608–1610 (2020).

44. De Wit, E., Van Doremalen, N., Falzarano, D. & Munster, V. J. SARS and MERS: Recent insights into emerging coronaviruses. Nat. Rev. Microbiol. 14, 523–534 (2016).

45. Clerici, M. et al. T-cell proliferation to subinfectious SIV correlates with lack of infection after challenge of macaques. Aids vol. 8 1391–1395 (1994).

46. English, K. M. et al. Contact among healthcare workers in the hospital setting: Developing the evidence base for innovative approaches to infection control. BMC Infect. Dis. 18, 1–12 (2018).

47. Chughtai, A. A. et al. Contamination by respiratory viruses on outer surface of medical masks used by hospital healthcare workers. BMC Infect. Dis. 19, 1–8 (2019).

48. Jacobs, J. L. et al. Use of surgical face masks to reduce the incidence of the common cold among health care workers in Japan: A randomized controlled trial. Am. J. Infect. Control 37, 417–419 (2009).

49. Sagar, M. et al. Recent endemic coronavirus infection is associated with less-severe COVID-19. J. Clin. Invest. 131, (2021).

50. Ng, K. W. et al. Preexisting and de novo humoral immunity to SARS-CoV-2 in humans. Science (80-.). 370, 1339–1343 (2020).

51. Yang, F. et al. Shared B cell memory to coronaviruses and other pathogens varies in human age groups and tissues. Science (80-.). 372, 738–741 (2021).

52. Wyllie, D. et al. SARS-CoV-2 responsive T cell numbers are associated with protection from COVID-19: A prospective cohort study in keyworkers. medRxiv (2020) doi:10.1101/2020.11.02.20222778.

53. Lipsitch, M., Grad, Y. H., Sette, A. & Crotty, S. Cross-reactive memory T cells and herd immunity to SARS-CoV-2. Nat. Rev. Immunol. 20, 709–713 (2020).

54. McMahan, K. et al. Correlates of protection against SARS-CoV-2 in rhesus macaques. Nature 590, 630–634 (2021).

55. Soresina, A. et al. Two X-linked agammaglobulinemia patients develop pneumonia as COVID-19 manifestation but recover. Pediatr. Allergy Immunol. 31, 565–569 (2020).

56. Mehta, P., Porter, J. C., Chambers, R. C., Isenberg, D. A. & Reddy, V. B-cell depletion with rituximab in the COVID-19 pandemic: where do we stand? Lancet Rheumatol. 2, e589–e590 (2020).

57. Cervia, C. et al. Systemic and mucosal antibody responses specific to SARS-CoV-2 during mild versus severe COVID-19. J. Allergy Clin. Immunol. 147, 545-557.e9 (2021).

## References (Methods)

58. Augusto, J. B. et al. Healthcare Workers Bioresource: Study outline and baseline characteristics of a prospective healthcare worker cohort to study immune protection and pathogenesis in COVID-19. Wellcome Open Res. 5, 179 (2020).

59. Reynolds, C. J. et al. Prior SARS-CoV-2 infection rescues B and T cell responses to variants after first vaccine dose. Science 5, 1–11 (2021).

60. O’Nions, J. et al. SARS-CoV-2 antibody responses in patients with acute leukaemia. Leukemia 35, 289–292 (2021).

61. Muir, L. et al. Neutralizing Antibody Responses After SARS-CoV-2 Infection in End-Stage Kidney Disease and Protection Against Reinfection. Kidney Int. Reports (2021) doi:10.1016/j.ekir.2021.03.902.

62. Capone, S. et al. Optimising T cell (re)boosting strategies for adenoviral and modified vaccinia Ankara vaccine regimens in humans. npj Vaccines 5, 1–14 (2020).

