## Supplementary Table 1 for "Pre-existing polymerase-specific T cells expand in abortive seronegative SARS-CoV-2 infection"

**Supplementary Table 1:** HCoV sequence accession numbers

| Accession | Viral species |
|---|---|
| MN369046 | Human coronavirus 229E |
| MN306046 | Human coronavirus 229E |
| MN306036 | Human coronavirus OC43 |
| MN306018 | Human coronavirus NL63 |
| MN310478 | Human coronavirus OC43 |
| MN310476 | Human coronavirus OC43 |
| MN306043 | Human coronavirus OC43 |
| MN306053 | Human coronavirus OC43 |
| MN306040 | Human coronavirus NL63 |
| MN306041 | Human coronavirus OC43 |
| MN306042 | Human coronavirus OC43 |
| MK334043 | Human coronavirus NL63 |
| MK334044 | Human coronavirus NL63 |
| MK334045 | Human coronavirus NL63 |
| MK334046 | Human coronavirus NL63 |
| MK334047 | Human coronavirus NL63 |
| MN026164 | Human coronavirus OC43 |
| MK167038 | Human coronavirus HKU1 |
| MH121121 | Human coronavirus OC43 |
| MH940245 | Human coronavirus HKU1 |
| MG197722 | Human coronavirus OC43 |
| MG197710 | Human coronavirus OC43 |
| MG197711 | Human coronavirus OC43 |
| MG197712 | Human coronavirus OC43 |
| MG197713 | Human coronavirus OC43 |
| MG197714 | Human coronavirus OC43 |
| MG197715 | Human coronavirus OC43 |
| MG197716 | Human coronavirus OC43 |
| MG197717 | Human coronavirus OC43 |
| MG197718 | Human coronavirus OC43 |
| MG197719 | Human coronavirus OC43 |
| MG197720 | Human coronavirus OC43 |
| MG197721 | Human coronavirus OC43 |
| MG197723 | Human coronavirus OC43 |
| MG197709 | Human coronavirus OC43 |
| MG977445 | Human coronavirus OC43 |
| MG977452 | Human coronavirus OC43 |
| MG977451 | Human coronavirus OC43 |
| MG977444 | Human coronavirus OC43 |
| MG977447 | Human coronavirus OC43 |
| MG977449 | Human coronavirus OC43 |
| MG428702 | Human coronavirus NL63 |
| MG428703 | Human coronavirus NL63 |
| MG428706 | Human coronavirus NL63 |
| MG428701 | Human coronavirus NL63 |
| MG428705 | Human coronavirus NL63 |
| MG428699 | Human coronavirus NL63 |
| MG428704 | Human coronavirus NL63 |
| MF314143 | Human coronavirus OC43 |
| MG772808 | Human coronavirus NL63 |
| KY014281 | Human coronavirus OC43 |
| KY014282 | Human coronavirus OC43 |
| MF374984 | Human coronavirus OC43 |
| MF374985 | Human coronavirus OC43 |
| MF374983 | Human coronavirus OC43 |
| MF542265 | Human coronavirus 229E |
| KY983587 | Human coronavirus 229E |
| KY983583 | Human coronavirus OC43 |
| KY983585 | Human coronavirus OC43 |
| KY983586 | Human coronavirus NL63 |
| KY983588 | Human coronavirus OC43 |
| KY967360 | Human coronavirus OC43 |
| KY621348 | Human coronavirus 229E |
| KY967359 | Human coronavirus OC43 |
| KY829118 | Human coronavirus NL63 |
| KY967357 | Human coronavirus 229E |
| KY967358 | Human coronavirus OC43 |
| KY967356 | Human coronavirus OC43 |
| KY967361 | Human coronavirus OC43 |
| KY554967 | Human coronavirus NL63 |
| KY554968 | Human coronavirus NL63 |
| KY554972 | Human coronavirus OC43 |
| KY554973 | Human coronavirus OC43 |
| KY554974 | Human coronavirus OC43 |
| KY554971 | Human coronavirus NL63 |
| KY554970 | Human coronavirus NL63 |
| KY554975 | Human coronavirus OC43 |
| KY674914 | Human coronavirus 229E |
| KY674919 | Human coronavirus 229E |
| KY674916 | Human coronavirus NL63 |
| KY674920 | Human coronavirus OC43 |
| KY674943 | Human coronavirus HKU1 |
| KY674921 | Human coronavirus HKU1 |
| KY684760 | Human coronavirus 229E |
| KY684759 | Human coronavirus OC43 |
| KX538965 | Human coronavirus OC43 |
| KX538966 | Human coronavirus OC43 |
| KX538967 | Human coronavirus OC43 |
| KX538968 | Human coronavirus OC43 |
| KX538969 | Human coronavirus OC43 |
| KX538970 | Human coronavirus OC43 |
| KX538971 | Human coronavirus OC43 |
| KX538972 | Human coronavirus OC43 |
| KX538973 | Human coronavirus OC43 |
| KX538974 | Human coronavirus OC43 |
| KX538976 | Human coronavirus OC43 |
| KX538979 | Human coronavirus OC43 |
| KX538975 | Human coronavirus OC43 |
| KX538977 | Human coronavirus OC43 |
| KX538978 | Human coronavirus OC43 |
| KX538964 | Human coronavirus OC43 |
| KY369905 | Human coronavirus OC43 |
| KY369906 | Human coronavirus OC43 |
| KY369907 | Human coronavirus OC43 |
| KY369913 | Human coronavirus 229E |
| KY369914 | Human coronavirus 229E |
| KY369909 | Human coronavirus 229E |
| KY369908 | Human coronavirus 229E |
| KY369911 | Human coronavirus 229E |
| KY369912 | Human coronavirus 229E |
| KX179500 | Human coronavirus NL63 |
| KX344031 | Human coronavirus OC43 |
| KU291448 | Human coronavirus 229E |
| KU521535 | Human coronavirus NL63 |
| KT779556 | Human coronavirus HKU1 |
| KT779555 | Human coronavirus HKU1 |
| KU131570 | Human coronavirus OC43 |
| KT381875 | Human coronavirus NL63 |
| KF923908 | Human coronavirus OC43 |
| KF923909 | Human coronavirus OC43 |
| KF923910 | Human coronavirus OC43 |
| KF923915 | Human coronavirus OC43 |
| KF923916 | Human coronavirus OC43 |
| KF923919 | Human coronavirus OC43 |
| KF923922 | Human coronavirus OC43 |
| KF923923 | Human coronavirus OC43 |
| KF923924 | Human coronavirus OC43 |
| KF923925 | Human coronavirus OC43 |
| KF923912 | Human coronavirus OC43 |
| KF923913 | Human coronavirus OC43 |
| KF923914 | Human coronavirus OC43 |
| KF923918 | Human coronavirus OC43 |
| KF923920 | Human coronavirus OC43 |
| KF923917 | Human coronavirus OC43 |
| KF923894 | Human coronavirus OC43 |
| KF923897 | Human coronavirus OC43 |
| KF923921 | Human coronavirus OC43 |
| KF923887 | Human coronavirus OC43 |
| KF923889 | Human coronavirus OC43 |
| KF923890 | Human coronavirus OC43 |
| KF923893 | Human coronavirus OC43 |
| KF923892 | Human coronavirus OC43 |
| KF923911 | Human coronavirus OC43 |
| KF923901 | Human coronavirus OC43 |
| KF923899 | Human coronavirus OC43 |
| KF923902 | Human coronavirus OC43 |
| KF923903 | Human coronavirus OC43 |
| KF923898 | Human coronavirus OC43 |
| KF923905 | Human coronavirus OC43 |
| KF923886 | Human coronavirus OC43 |
| KF923904 | Human coronavirus OC43 |
| KF923907 | Human coronavirus OC43 |
| KF923895 | Human coronavirus OC43 |
| KT266906 | Human coronavirus NL63 |
| KF530110 | Human coronavirus NL63 |
| KF530104 | Human coronavirus NL63 |
| KF530105 | Human coronavirus NL63 |
| KF530107 | Human coronavirus NL63 |
| KF530108 | Human coronavirus NL63 |
| KF530114 | Human coronavirus NL63 |
| KF530106 | Human coronavirus NL63 |
| KF530113 | Human coronavirus NL63 |
| KF530109 | Human coronavirus NL63 |
| KF530111 | Human coronavirus NL63 |
| KF530112 | Human coronavirus NL63 |
| KF686340 | Human coronavirus HKU1 |
| KF686342 | Human coronavirus HKU1 |
| KF686343 | Human coronavirus HKU1 |
| KF686344 | Human coronavirus HKU1 |
| KF686341 | Human coronavirus HKU1 |
| KF686346 | Human coronavirus HKU1 |
| KF514430 | Human coronavirus 229E |
| KF514433 | Human coronavirus 229E |
| KF514432 | Human coronavirus 229E |
| KF430201 | Human coronavirus HKU1 |
| JX504050 | Human coronavirus NL63 |
| JX503061 | Human coronavirus 229E |
| JX503060 | Human coronavirus 229E |
| JX524171 | Human coronavirus NL63 |
| JX104161 | Human coronavirus NL63 |
| JQ765571 | Human coronavirus NL63 |
| JQ765564 | Human coronavirus NL63 |
| JQ765572 | Human coronavirus NL63 |
| JQ765568 | Human coronavirus NL63 |
| JQ765570 | Human coronavirus NL63 |
| JQ765574 | Human coronavirus NL63 |
| JQ765565 | Human coronavirus NL63 |
| JQ765567 | Human coronavirus NL63 |
| JQ765573 | Human coronavirus NL63 |
| JQ765563 | Human coronavirus NL63 |
| JQ765575 | Human coronavirus NL63 |
| JQ765566 | Human coronavirus NL63 |
| JQ765569 | Human coronavirus NL63 |
| HM034837 | Human coronavirus HKU1 |
| DQ445911 | Human coronavirus NL63 |
| DQ445912 | Human coronavirus NL63 |
| DQ415896 | Human coronavirus HKU1 |
| DQ415897 | Human coronavirus HKU1 |
| DQ415898 | Human coronavirus HKU1 |
| DQ415899 | Human coronavirus HKU1 |
| DQ415900 | Human coronavirus HKU1 |
| DQ415901 | Human coronavirus HKU1 |
| DQ415902 | Human coronavirus HKU1 |
| DQ415903 | Human coronavirus HKU1 |
| DQ415904 | Human coronavirus HKU1 |
| DQ415905 | Human coronavirus HKU1 |
| DQ415906 | Human coronavirus HKU1 |
| DQ415907 | Human coronavirus HKU1 |
| DQ415908 | Human coronavirus HKU1 |
| DQ415909 | Human coronavirus HKU1 |
| DQ415910 | Human coronavirus HKU1 |
| DQ415911 | Human coronavirus HKU1 |
| DQ415912 | Human coronavirus HKU1 |
| DQ415913 | Human coronavirus HKU1 |
| DQ415914 | Human coronavirus HKU1 |
| AY597011 | Human coronavirus HKU1 |
