## Supplementary Table 2 for "Pre-existing polymerase-specific T cells expand in abortive seronegative SARS-CoV-2 infection"

**Supplementary Table 2:** 2D epitope mapping matrices.

| NSP7 | Pool A | Pool B | Pool C | Pool D |
|---|---|---|---|---|
| Pool E | 7-1 | 7-2 | 7-3 | 7-4 |
| Pool F | 7-5 | 7-6 | 7-7 | 7-8 |
| Pool G | 7-9 | 7-10 | 7-11 | 7-12 |
| Pool H | 7-13 | 7-14 | 7-15 | x |

| NSP12-2 | Pool A | Pool B | Pool C | Pool D | Pool E | Pool F | |
|---|---|---|---|---|---|---|---|
| Pool G | 12-38 | 12-39 | 12-40 | 12-41 | 12-42 | 12-43 | x |
| Pool H | 12-44 | 12-45 | 12-46 | 12-47 | 12-48 | 12-49 | x |
| Pool I | 12-50 | 12-51 | 12-52 | 12-53 | 12-54 | 12-55 | x |
| Pool J | 12-56 | 12-57 | 12-58 | 12-59 | 12-60 | 12-61 | x |
| Pool K | 12-62 | 12-63 | 12-64 | 12-65 | 12-66 | 12-67 | x |
| Pool L | 12-68 | 12-69 | 12-70 | 12-71 | 12-72 | 12-73 | 12-74 |

| NSP12-3 | Pool A | Pool B | Pool C | Pool D | Pool E | Pool F | |
|---|---|---|---|---|---|---|---|
| Pool G | 12-75 | 12-76 | 12-77 | 12-78 | 12-79 | 12-80 | x |
| Pool H | 12-81 | 12-82 | 12-83 | 12-84 | 12-85 | 12-86 | x |
| Pool I | 12-87 | 12-88 | 12-89 | 12-90 | 12-91 | 12-92 | x |
| Pool J | 12-93 | 12-94 | 12-95 | 12-96 | 12-97 | 12-98 | x |
| Pool K | 12-99 | 12-100 | 12-101 | 12-102 | 12-103 | 12-104 | x |
| Pool L | 12-105 | 12-106 | 12-107 | 12-108 | 12-109 | 12-110 | 12-111 |

| NSP12-5 | Pool A | Pool B | Pool C | Pool D | Pool E | Pool F | |
|---|---|---|---|---|---|---|---|
| Pool G | 12-149 | 12-150 | 12-151 | 12-152 | 12-153 | 12-154 | x |
| Pool H | 12-155 | 12-156 | 12-157 | 12-158 | 12-159 | 12-160 | x |
| Pool I | 12-161 | 12-162 | 12-163 | 12-164 | 12-165 | 12-166 | x |
| Pool J | 12-167 | 12-168 | 12-169 | 12-170 | 12-171 | 12-172 | x |
| Pool K | 12-173 | 12-174 | 12-175 | 12-176 | 12-177 | 12-178 | x |
| Pool L | 12-179 | 12-180 | 12-181 | 12-182 | 12-183 | 12-184 | 12-185 |

| NSP13-3 | Pool A | Pool B | Pool C | Pool D | Pool E | Pool F | PoolG |
|---|---|---|---|---|---|---|---|
| Pool H | 13-79 | 13-80 | 13-81 | 13-82 | 13-83 | 13-84 | 13-85 |
| Pool I | 13-86 | 13-87 | 13-88 | 13-89 | 13-90 | 13-91 | 13-92 |
| Pool J | 13-93 | 13-94 | 13-95 | 13-96 | 13-97 | 13-98 | 13-99 |
| Pool K | 13-100 | 13-101 | 13-102 | 13-103 | 13-104 | 13-105 | 13-106 |
| Pool L | 13-107 | 13-108 | 13-109 | 13-110 | 13-111 | 13-112 | 13-113 |
| Pool M | 13-114 | 13-115 | 13-116 | 13-117 | 13-118 | x | x |
