## Supplementary Table 3 for "Pre-existing polymerase-specific T cells expand in abortive seronegative SARS-CoV-2 infection"

**Supplementary Table 3:** NSP12 Overlapping peptide sequences.

### SARS-Cov Peptides NSP 12

| # peptide | NSP12 | aa |
|---|---|---|
| 12-1 | SADASTFLNRVCGVS | 1-15 |
| 12-2 | TFLNRVCGVSAARLT | 6-20 |
| 12-3 | VCGVSAARLTPCGTG | 11-25 |
| 12-4 | AARLTPCGTGTSTDV | 16-30 |
| 12-5 | PCGTGTSTDVVYRAF | 21-35 |
| 12-6 | TSTDVVYRAFDIYNE | 26-40 |
| 12-7 | VYRAFDIYNEKVAGF | 31-45 |
| 12-8 | DIYNEKVAGFAKFLK | 36-50 |
| 12-9 | KVAGFAKFLKTNCCR | 41-55 |
| 12-10 | AKFLKTNCCRFQEKD | 46-60 |
| 12-11 | TNCCRFQEKDEEGNL | 51-65 |
| 12-12 | FQEKDEEGNLLDSYF | 56-70 |
| 12-13 | EEGNLLDSYFVVKRH | 61-75 |
| 12-14 | LDSYFVVKRHTMSNY | 66-80 |
| 12-15 | VVKRHTMSNYQHEET | 71-85 |
| 12-16 | TMSNYQHEETIYNLV | 76-90 |
| 12-17 | QHEETIYNLVKDCPA | 81-95 |
| 12-18 | IYNLVKDCPAVAVHD | 86-100 |
| 12-19 | KDCPAVAVHDFFKFR | 91-105 |
| 12-20 | VAVHDFFKFRVDGDM | 96-110 |
| 12-21 | FFKFRVDGDMVPHIS | 101-115 |
| 12-22 | VDGDMVPHISRQRLT | 106-120 |
| 12-23 | VPHISRQRLTKYTMA | 111-125 |
| 12-24 | RQRLTKYTMADLVYA | 116-130 |
| 12-25 | KYTMADLVYALRHFD | 121-135 |
| 12-26 | DLVYALRHFDEGNCD | 126-140 |
| 12-27 | LRHFDEGNCDTLKEI | 131-145 |
| 12-28 | EGNCDTLKEILVTYN | 136-150 |
| 12-29 | TLKEILVTYNCCDDD | 141-155 |
| 12-30 | LVTYNCCDDDYFNKK | 146-160 |
| 12-31 | CCDDDYFNKKDWYDF | 151-165 |
| 12-32 | YFNKKDWYDFVENPD | 156-170 |
| 12-33 | DWYDFVENPDILRVY | 161-175 |
| 12-34 | VENPDILRVYANLGE | 166-180 |
| 12-35 | ILRVYANLGERVRQS | 171-185 |
| 12-36 | ANLGERVRQSLLKTV | 176-190 |
| 12-37 | RVRQSLLKTVQFCDA | 181-195 |
| 12-38 | LLKTVQFCDAMRDAG | 186-200 |
| 12-39 | QFCDAMRDAGIVGVL | 191-205 |
| 12-40 | MRDAGIVGVLTLDNQ | 196-210 |
| 12-41 | IVGVLTLDNQDLNGN | 201-215 |
| 12-42 | TLDNQDLNGNWYDFG | 206-220 |
| 12-43 | DLNGNWYDFGDFVQV | 211-225 |
| 12-44 | WYDFGDFVQVAPGCG | 216-230 |
| 12-45 | DFVQVAPGCGVPIVD | 221-235 |
| 12-46 | APGCGVPIVDSYYSL | 226-240 |
| 12-47 | VPIVDSYYSLLMPIL | 231-245 |
| 12-48 | SYYSLLMPILTLTRA | 236-250 |
| 12-49 | LMPILTLTRALAAES | 241-255 |
| 12-50 | TLTRALAAESHMDAD | 246-260 |
| 12-51 | LAAESHMDADLAKPL | 251-265 |
| 12-52 | HMDADLAKPLIKWDL | 256-270 |
| 12-53 | LAKPLIKWDLLKYDF | 261-275 |
| 12-54 | IKWDLLKYDFTEERL | 266-280 |
| 12-55 | LKYDFTEERLCLFDR | 271-285 |
| 12-56 | TEERLCLFDRYFKYW | 276-290 |
| 12-57 | CLFDRYFKYWDQTYH | 281-295 |
| 12-58 | YFKYWDQTYHPNCIN | 286-300 |
| 12-59 | DQTYHPNCINCLDDR | 291-305 |
| 12-60 | PNCINCLDDRCILHC | 296-310 |
| 12-61 | CLDDRCILHCANFNV | 301-315 |
| 12-62 | CILHCANFNVLFSTV | 306-320 |
| 12-63 | ANFNVLFSTVFPPTS | 311-325 |
| 12-64 | LFSTVFPPTSFGPLV | 316-330 |
| 12-65 | FPPTSFGPLVRKIFV | 321-335 |
| 12-66 | FGPLVRKIFVDGVPF | 326-340 |
| 12-67 | RKIFVDGVPFVVSTG | 331-345 |
| 12-68 | DGVPFVVSTGYHFRE | 336-350 |
| 12-69 | VVSTGYHFRELGVVH | 341-355 |
| 12-70 | YHFRELGVVHNQDVN | 346-360 |
| 12-71 | LGVVHNQDVNLHSSR | 351-365 |
| 12-72 | NQDVNLHSSRLSFKE | 356-370 |
| 12-73 | LHSSRLSFKELLVYA | 361-375 |
| 12-74 | LSFKELLVYAADPAM | 366-380 |
| 12-75 | LLVYAADPAMHAASG | 371-385 |
| 12-76 | ADPAMHAASGNLLLD | 376-390 |
| 12-77 | HAASGNLLLDKRTTC | 381-395 |
| 12-78 | NLLLDKRTTCFSVAA | 386-400 |
| 12-79 | KRTTCFSVAALTNNV | 391-405 |
| 12-80 | FSVAALTNNVAFQTV | 396-410 |
| 12-81 | LTNNVAFQTVKPGNF | 401-415 |
| 12-82 | AFQTVKPGNFNKDFY | 406-420 |
| 12-83 | KPGNFNKDFYDFAVS | 411-425 |
| 12-84 | NKDFYDFAVSKGFFK | 416-430 |
| 12-85 | DFAVSKGFFKEGSSV | 421-435 |
| 12-86 | KGFFKEGSSVELKHF | 426-440 |
| 12-87 | EGSSVELKHFFFAQD | 431-445 |
| 12-88 | ELKHFFFAQDGNAAI | 436-450 |
| 12-89 | FFAQDGNAAISDYDY | 441-455 |
| 12-90 | GNAAISDYDYYRYNL | 446-460 |
| 12-91 | SDYDYYRYNLPTMCD | 451-465 |
| 12-92 | YRYNLPTMCDIRQLL | 456-470 |

### SARS-Cov-2 Peptides NSP 12

Divergence in sequence from SARS-CoV to SARS-CoV-2 are highlighted in bold

**NSP12-2**

| # peptide | SARS-CoV-2 NSP12 | aa |
|---|---|---|
| 12-38 | LLKTVQFCDAMR**N**AG | 186-200 |
| 12-39 | QFCDAMR**N**AGIVGVL | 191-205 |
| 12-40 | MR**N**AGIVGVLTLDNQ | 196-210 |
| 12-43 | DLNGNWYDFGDF**I**Q**T** | 211-225 |
| 12-44 | WYDFGDF**I**Q**TT**PG**S**G | 216-230 |
| 12-45 | DF**I**Q**TT**PG**S**GVPVVD | 221-235 |
| 12-46 | **T**PG**S**GVP**V**VDSYYSL | 226-240 |
| 12-47 | VP**V**VDSYYSLLMPIL | 231-245 |
| 12-49 | LMPILTLTRAL**T**AES | 241-255 |
| 12-50 | TLTRAL**T**AESH**V**D**T**D | 246-260 |
| 12-51 | L**T**AESH**V**D**T**DL**T**KP**Y** | 251-265 |
| 12-52 | H**V**D**T**DL**T**KP**Y**IKWDL | 256-270 |
| 12-53 | L**T**KP**Y**IKWDLLKYDF | 261-275 |
| 12-55 | LKYDFTEERL**K**LFDR | 271-285 |
| 12-56 | TEERL**K**LFDRYFKYW | 276-290 |
| 12-57 | **K**LFDRYFKYWDQTYH | 281-295 |
| 12-58 | YFKYWDQTYHPNC**V**N | 286-300 |
| 12-59 | DQTYHPNC**V**NCLDDR | 291-305 |
| 12-60 | PNC**V**NCLDDRCILHC | 296-310 |

| | | |
|---|---|---|
| 12-93 | PTMCDIRQLLFVVEV | 461-475 |
| 12-94 | IRQLLFVVEVVDKYF | 466-480 |
| 12-95 | FVVEVVDKYFDCYDG | 471-485 |
| 12-96 | VDKYFDCYDGGCINA | 476-490 |
| 12-97 | DCYDGGCINANQVIV | 481-495 |
| 12-98 | GCINANQVIVNNLDK | 486-500 |
| 12-99 | NQVIVNNLDKSAGFP | 491-505 |
| 12-100 | NNLDKSAGFPFNKWG | 496-510 |
| 12-101 | SAGFPFNKWGKARLY | 501-515 |
| 12-102 | FNKWGKARLYYDSMS | 506-520 |
| 12-103 | KARLYYDSMSYEDQD | 511-525 |
| 12-104 | YDSMSYEDQDALFAY | 516-530 |
| 12-105 | YEDQDALFAYTKRNV | 521-535 |
| 12-106 | ALFAYTKRNVIPTIT | 526-540 |
| 12-107 | TKRNVIPTITQMNLK | 531-545 |
| 12-108 | IPTITQMNLKYAISA | 536-550 |
| 12-109 | QMNLKYAISAKNRAR | 541-555 |
| 12-110 | YAISAKNRARTVAGV | 546-560 |
| 12-111 | KNRARTVAGVSICST | 551-565 |
| 12-112 | TVAGVSICSTMTNRQ | 556-570 |
| 12-113 | SICSTMTNRQFHQKL | 561-575 |
| 12-114 | MTNRQFHQKLLKSIA | 566-580 |
| 12-115 | FHQKLLKSIAATRGA | 571-585 |
| 12-116 | LKSIAATRGATVVIG | 576-590 |
| 12-117 | ATRGATVVIGTSKFY | 581-595 |
| 12-118 | TVVIGTSKFYGGWHN | 586-600 |
| 12-119 | TSKFYGGWHNMLKTV | 591-605 |
| 12-120 | GGWHNMLKTVYSDVE | 596-610 |
| 12-121 | MLKTVYSDVETPHLM | 601-615 |
| 12-122 | YSDVETPHLMGWDYP | 606-620 |
| 12-123 | TPHLMGWDYPKCDRA | 611-625 |
| 12-124 | GWDYPKCDRAMPNML | 616-630 |
| 12-125 | KCDRAMPNMLRIMAS | 621-635 |
| 12-126 | MPNMLRIMASLVLAR | 626-640 |
| 12-127 | RIMASLVLARKHNTC | 631-645 |
| 12-128 | LVLARKHNTCCNLSH | 636-650 |
| 12-129 | KHNTCCNLSHRFYRL | 641-655 |
| 12-130 | CNLSHRFYRLANECA | 646-660 |
| 12-131 | RFYRLANECAQVLSE | 651-665 |
| 12-132 | ANECAQVLSEMVMCG | 656-670 |
| 12-133 | QVLSEMVMCGGSLYV | 661-675 |
| 12-134 | MVMCGGSLYVKPGGT | 666-680 |
| 12-135 | GSLYVKPGGTSSGDA | 671-685 |
| 12-136 | KPGGTSSGDATTAYA | 676-690 |
| 12-137 | SSGDATTAYANSVFN | 681-695 |
| 12-138 | TTAYANSVFNICQAV | 686-700 |
| 12-139 | NSVFNICQAVTANVN | 691-705 |
| 12-140 | ICQAVTANVNALLST | 696-710 |
| 12-141 | TANVNALLSTDGNKI | 701-715 |
| 12-142 | ALLSTDGNKIADKYV | 706-720 |
| 12-143 | DGNKIADKYVRNLQH | 711-725 |
| 12-144 | ADKYVRNLQHRLYEC | 716-730 |
| 12-145 | RNLQHRLYECLYRNR | 721-735 |
| 12-146 | RLYECLYRNRDVDHE | 726-740 |
| 12-147 | LYRNRDVDHEFVDEF | 731-745 |
| 12-148 | DVDHEFVDEFYAYLR | 736-750 |
| 12-149 | FVDEFYAYLRKHFSM | 741-755 |
| 12-150 | YAYLRKHFSMMILSD | 746-760 |
| 12-151 | KHFSMMILSDDAVVC | 751-765 |
| 12-152 | MILSDDAVVCYNSNY | 756-770 |
| 12-153 | DAVVCYNSNYAAQGL | 761-775 |
| 12-154 | YNSNYAAQGLVASIK | 766-780 |
| 12-155 | AAQGLVASIKNFKAV | 771-785 |
| 12-156 | VASIKNFKAVLYYQN | 776-790 |
| 12-157 | NFKAVLYYQNNVFMS | 781-795 |
| 12-158 | LYYQNNVFMSEAKCW | 786-800 |
| 12-159 | NVFMSEAKCWTETDL | 791-805 |
| 12-160 | EAKCWTETDLTKGPH | 796-810 |
| 12-161 | TETDLTKGPHEFCSQ | 801-815 |
| 12-162 | TKGPHEFCSQHTMLV | 806-820 |
| 12-163 | EFCSQHTMLVKQGDD | 811-825 |
| 12-164 | HTMLVKQGDDYVYLP | 816-830 |
| 12-165 | KQGDDYVYLPYPDPS | 821-835 |
| 12-166 | YVYLPYPDPSRILGA | 826-840 |
| 12-167 | YPDPSRILGAGCFVD | 831-845 |
| 12-168 | RILGAGCFVDDIVKT | 836-850 |
| 12-169 | GCFVDDIVKTDGTLM | 841-855 |
| 12-170 | DIVKTDGTLMIERFV | 846-860 |
| 12-171 | DGTLMIERFVSLAID | 851-865 |
| 12-172 | IERFVSLAIDAYPLT | 856-870 |
| 12-173 | SLAIDAYPLTKHPNQ | 861-875 |
| 12-174 | AYPLTKHPNQEYADV | 866-880 |
| 12-175 | KHPNQEYADVFHLYL | 871-885 |
| 12-176 | EYADVFHLYLQYIRK | 876-890 |
| 12-177 | FHLYLQYIRKLHDEL | 881-895 |
| 12-178 | QYIRKLHDELTGHML | 886-900 |
| 12-179 | LHDELTGHMLDMYSV | 891-905 |
| 12-180 | TGHMLDMYSVMLTND | 896-910 |
| 12-181 | DMYSVMLTNDNTSRY | 901-915 |
| 12-182 | MLTNDNTSRYWEPEF | 906-920 |
| 12-183 | NTSRYWEPEFYEAMY | 911-925 |
| 12-184 | WEPEFYEAMYTPHTV | 916-930 |
| 12-185 | PEFYEAMYTPHTVLQ | 921-935 |

| | | | |
|---|---|---|---|
| **NSP12-5** | 12-149 | FV**N**EFYAYLRKHFSM | 741-755 |
| | 12-152 | MILSDDAVVC**F**NS**T**Y | 756-770 |
| | 12-153 | DAVVC**F**NS**T**YA**S**QGL | 761-775 |
| | 12-154 | **F**NS**T**YA**S**QGLVASIK | 766-780 |
| | 12-155 | A**S**QGLVASIKNFK**S**V | 771-785 |
| | 12-156 | VASIKNFK**S**VLYYQN | 776-790 |
| | 12-157 | NFK**S**VLYYQNNVFMS | 781-795 |
